# PCSK6 and Survival in Idiopathic Pulmonary Fibrosis

**DOI:** 10.1101/2022.05.06.22274705

**Authors:** Justin M. Oldham, Richard J. Allen, Jose M. Lorenzo-Salazar, Philip L. Molyneaux, Shwu-Fan Ma, Chitra Joseph, John S. Kim, Beatriz Guillen-Guio, Tamara Hernández-Beeftink, Jonathan A. Kropski, Yong Huang, Cathryn T. Lee, Ayodeji Adegunsoye, Janelle Vu Pugashetti, Angela L. Linderholm, Vivian Vo, Mary E. Strek, Jonathan Jou, Adrian Muñoz-Barrera, Luis A. Rubio-Rodriguez, Richard Hubbard, Nik Hirani, Moira K.B. Whyte, Simon Hart, Andrew G. Nicholson, Lisa Lancaster, Helen Parfrey, Doris Rassl, William Wallace, Eleanor Valenzi, Yingze Zhang, Josyf Mychaleckyj, Amy Stockwell, Naftali Kaminski, Paul J. Wolters, Maria Molina-Molina, William A. Fahy, Fernando J. Martinez, Ian P. Hall, Martin D. Tobin, Toby M. Maher, Timothy S. Blackwell, Brian L. Yaspan, R Gisli Jenkins, Carlos Flores, Louise V Wain, Imre Noth

**Author notes:** These authors contributed equally (Justin M. Oldham, Richard J. Allen, Jose M. Lorenzo-Salazar, Philip L. Molyneaux). These authors contributed equally (R Gisli Jenkins, Carlos Flores, Louise V Wain, Imre Noth). **Corresponding Author** Justin M. Oldham, 6301 MSRB3, 1150 W. Medical Center Dr., Ann Arbor, MI 48109-5642. **Author Contributions** JMO, RJA, JMLS, SFM, RGJ, CF, LVW and IN designed and supervised the study. JMO, PLM, JAK, AA, JVP, MES, RH, NH, MW, SH, HP, NK, PW, MMM, FJM, TMM, TSB, RGJ and IN recruited patients for the study. JMO, PLM, SFM, JAK, AL, VV, YZ, AS, NK, PW, IH, MDT, WAF, TMM, TSB, BLY, RGJ, CF and IN contributed to DNA extraction and genotyping. CJ, AGN, DR, WW and RGJ secured lung tissue and performed the IHC analysis. JMO, RJA, JMLS, CJ, JSK, BGG, THB, YH, AS, BLY, RGJ, CF, LVW performed the analyses. JMO, PLM, JK, CTL, AA, JVP, MES, EV, YZ, KG, DK, NK, PW, MMM, FJM, TMM, RGJ and IN collected paired clinical data. JMO, JMLS, RJA, RGJ, CF, LW and IN wrote the manuscript with the input of all authors. All authors approved the final version of this manuscript. **Funding** NHLBI - K23HL138190 (JMO), R56HL158935 (JMO), K23HL150301 (JSK), K23HL146942 (AA), T32HL007605 (CTL), RO1HL130796 (IN), UG3HL145266 (IN). Ministry of Science and Innovation (RTC-2017-6471-1; AEI/FEDER UE (CF). Instituto de Salud Carlos III co-financed by the ERDF “A way of making Europe” from the EU (PI20/00876, CF), Instituto Tecnológico y de Energías Renovables (OA17/008 (CF) Consejería de Educación-Gobierno de Canarias and Cabildo Insular de Tenerife (BOC n.º 173, 24/08/2017 (JMLS). British Lung Foundation Chair in Respiratory Research (C17-3) (TMM), GlaxoSmithKline/British Lung Foundation Chair in Respiratory Research (C17-1) (LWW) Wellcome Trust - WT202849/Z/16/Z (MDT), 221680/Z/20/Z (BGG). For the purpose of open access, the author has applied a CC BY public copyright license to any Author Accepted Manuscript version arising from this submission. BREATHE — The Health Data Research Hub for Respiratory Health (MC_PC_19004) (LWW) Action for Pulmonary Fibrosis Research Fellows (RJA and PLM). This research used the ALICE and SPECTRE High Performance Computing Facilities at the University of Leicester and TeideHPC (https://teidehpc.iter.es/en/home) at Instituto Tecnológico y de Energías Renovables. **Role of the Funding Source** This study was funded by the agencies listed above, which had no role in the design or conduct of the study. All authors had full access to all data, and the corresponding author had final responsibility for the decision to submit for publication.

## Abstract

**Rationale:** Idiopathic pulmonary fibrosis (IPF) is a devastating disease characterized by limited treatment options and high mortality. Novel therapies and prognostic biomarkers are needed.

**Objective:** To identify and validate molecular determinants of IPF survival.

**Methods:** A staged genome-wide association study (GWAS) was performed using paired genomic and survival data. Stage I cases were drawn from centers across the US and Europe and stage II cases from Vanderbilt University. Cox proportional hazards regression was used to identify gene variants associated with differential transplant-free survival (TFS). Stage I variants with nominal significance (*p*<5×10^−5^) were advanced for stage II testing and meta-analyzed to identify those reaching genome-wide significance (*p*<5×10^−8^). Downstream analyses were performed for genes and proteins associated with variants reaching genome-wide significance.

**Main Results:** After quality controls, 1481 stage I cases and 397 stage II cases were included in the analysis. After filtering, 9,075,629 variants were tested in stage I, with 158 meeting advancement criteria. Four variants associated with TFS with consistent effect direction were identified in stage II, including one in an intron of proprotein convertase subtilisin/kexin type 6 (*PCSK6*) reaching genome-wide significance (HR 4.11; 95%CI 2.54-6.67; *p*=9.45×10^−9^). PCSK6 protein was highly expressed in IPF lung parenchyma and negatively correlated with survival. Peripheral blood *PCSK6* gene expression and plasma concentration were associated with reduced transplant-free survival.

**Conclusions:** We identified four novel variants associated with IPF survival, including one in *PCSK6* that reached genome-wide significance. Downstream analyses suggested that PCSK6 protein may serve as prognostic biomarker in IPF and potential therapeutic target.

## Introduction

Idiopathic Pulmonary Fibrosis (IPF) is a devastating disease characterized by progressive lung scarring and poor survival.^1,2^ Two anti-fibrotic therapies have been approved for the treatment of IPF after randomized controlled trials demonstrated efficacy in slowing lung function decline.^3,4^ Despite this advance, outcomes remain poor and anti-fibrotic therapy appears to provide only modest survival benefit.^5^ To improve IPF outcomes, novel therapeutic targets are needed.

We and others have identified molecular IPF risk factors through unbiased investigation of the genome, transcriptome, and proteome.^6-15^ Among the strongest molecular determinants of IPF is a common variant in the promoter region of *MUC5B*, which increases the odds of developing IPF by nearly 5-fold per risk allele.^6-9^ Despite this strong association with IPF onset, the MUC5B promoter was paradoxically associated with improved survival.^16^ Few other susceptibility-associated gene variants have been shown to reliably predict differential IPF survival, suggesting that molecular determinants of IPF susceptibility and progression may have limited overlap.

To better understand molecular drivers of IPF progression and identify new therapeutic targets, we conducted a two-stage, multi-center, international genome-wide association study (GWAS) of IPF survival, followed by downstream analysis of genes and proteins associated with top survival-associated variant to determine whether these circulating biomarkers also predicted differential survival.

## Methods

### Cohorts and case selection

All patients provided informed consent for research blood draw in accordance with protocols approved by the institutional review board at each participating institution. GWAS stage I cases consisted of unrelated IPF patients of European ancestry from three previously described case-control GWAS datasets from the United States (US),^9^ United Kingdom (UK),^7^ and a combined cohort from the US, UK and Spain (UUS)^6^. Available outcome data was gathered for all cases meeting international consensus criteria for IPF^17^ and survival plotted for individual cohorts within each dataset. Patients without available outcome data were excluded, as were clinical trial cohorts due to short follow-up (**Supplemental Methods**). Stage II cases consisted of previously described, unrelated IPF patients of European ancestry from Vanderbilt University.^18^

### Genotyping and quality control

Genotypes were generated for stage I cases using SNP (single nucleotide polymorphism) genotyping arrays (Thermo Fisher Scientific, Waltham, MA) according to previously described methods.^6,7,9^ Imputation for stage I cases was performed using the Michigan Imputation Server using the Haplotype Reference Consortium panel (v1.1 2016). Genotypes for the stage II cases were determined by whole genome sequencing, as previously described.^18^ Stringent quality control measures were applied with a two-tier variant filtering scheme as follows: those ranging 0.5≤MAF<1% were retained when imputation R^2^≥0.8; and those with MAF≥1% when imputation R^2^ ≥ 0.5. In addition, variants deviating from Hardy-Weinberg equilibrium (*p*<1.0×10^−6^) were removed.

### Genome-wide survival analysis

The primary endpoint assessed was transplant-free survival (TFS), defined as the time in months from site-determined date of IPF diagnosis to event (death or lung transplant) or censoring date. Variants associated with differential TFS were identified using a multivariable Cox proportional hazards regression model adjusted for age, sex, center, and first ten genetic principal components. Variant genotypes were treated as a continuous variable with each patient having an imputed genotype dosage between zero and two risk alleles. Variants present in at least two datasets (US, UK and UUS) were meta-analyzed using METAL (v2011-03-25) to generate stage I results.

Variants nominally associated with TFS in stage I were defined as those with Wald *p*<0.05 in at least two datasets with the same direction of effect and p<5.0×10^−5^ in stage I meta-analysis. Conditional analysis of these SNPs to deduce their independence was performed with GCTA-COJO v1.26. The proportional hazards assumption was then assessed for each independent variant meeting advancement criteria by testing whether Schoenfeld residual rank varied by genotype strata. Variants that satisfied the proportional hazards assumption were advanced for stage II testing. Stage I and II cases were then meta-analyzed using METAL with the genome-wide significance threshold set at *p*<5.0×10^−8^. *In silico* assessments were used to infer the biological effect of variants associated with TFS after stage II testing.

### PCSK6 tissue expression

Formalin-fixed paraffin-embedded human lung tissue sections obtained from patients with IPF undergoing surgical lung biopsy were compared to control subjects undergoing lung resection for malignancy, with sections distal to areas of malignancy utilized.Immunohistochemistry was performed using standard methods (**Supplemental Methods**) and mean staining intensity of PCSK6 protein was compared between IPF cases and non-IPF controls using a Mann-Whitney U-test.

### PCSK6 clinical outcome association

The association between circulating *PCSK6* gene expression and TFS was assessed using three previously published microarray datasets from the COMET trial, Imperial College and the University of Chicago, which were analyzed separately with results meta-analyzed and presented as a forest plot.^19^ Circulating plasma PCSK6 protein concentration was then determined in patients with IPF from UC-Davis and UChicago (**Supplemental Methods**), log^2^ transformed, and tested for TFS association using Cox proportional hazards regression.^20^ The proportional hazards assumption was satisfied for all downstream survival analyses.

## Results

### Case selection for stage I

Patients comprising the US cohort included those from the University of Chicago (n=118) and University of Pittsburgh (n=200) **(Figure E1, Figure E2)**. Those comprising the UK cohort included patients from the University of Edinburgh (n=119), Trent Lung Fibrosis Study (n=210), a subset of those participating in the prospective, multi-center PROFILE study (NCT 01134822) (n=175), and aggregated patients from smaller UK centers (Hull and Papworth) (n=61) **(Figure E1, Figure E3)**. Patients comprising the UUS cohort included those from the University of Chicago (independent of those from the US cohort, n=187), PROFILE study (independent of those in the UK dataset, n=299), University of California (Davis and San Francisco) (n=84) and aggregated patients from centers in Spain (n=28) **(Figure E1, Figure E4)**.

### Baseline characteristics and outcomes

Following phenotypic exclusions, 1481 patients comprising stage I were included in the analysis. These included 318 patients from the US dataset, 565 from the UK dataset, and 598 from the UUS dataset. Baseline characteristics for each dataset are shown in **Table 1**. The mean age ranged from 67 to 72 years and males comprised 71-75% of each dataset. The mean percent predicted FVC and diffusion capacity of the lung for carbon monoxide (DLCO) was lowest in patients comprising the US dataset and highest in the UK dataset. A majority of patients in each dataset were classified as gender, age, physiology^21^ (GAP) stage I or II. Death events were highest in the UK cohort and lowest in the UUS cohort (**Figure E5**). Median survival was 39.3 months in the US dataset, 53.2 months in the UK dataset and 40.6 months in the UUS dataset. Median survival was 48 months in the Vanderbilt University validation cohort (**Figure E6**).

**Table 1.** Baseline characteristics and outcomes of stage I and II datasets.

| **Characteristic** | **Stage I (n=1481)** |  |  | **Stage II (n=397)** |
| --- | --- | --- | --- | --- |
|  | **US\* (n=318)** | **UK\*\* (n=565)** | **UUS\*\*\* (n=598)** | **Vanderbilt\*\*\*\*** |
| Age | 67.3 (8.9) | 72.1 (8.4) | 69.8 (8.3) | 65.7 (9.0) |
| Male sex | 234 (73.6) | 403 (71.3) | 449 (75.1) | 295 (74.3) |
| FVC, % predicted | 62.7 (17.9) | 77.7 (18.5) | 70.2 (18.6) | 65.0 (16.1) |
| DLCO, % predicted | 36.7 (14.6) | 43.3 (14.3) | 39.9 (14.6) | 39.1 (13.5) |
| GAP Stage |  |  |  |  |
| I | 72 (23.1) | 106 (35.2) | 135 (24.2) | 115 (29.8) |
| II | 149 (47.8) | 144 (47.8) | 286 (51.4) | 201 (52.1) |
| III | 91 (29.2) | 51 (16.9) | 136 (24.4) | 70 (18.1) |
| Death | 189 (59.4) | 366 (64.8) | 257 (43.0) | 202 (50.9) |
| Transplant | 52 (16.4) | 2 (0.4) | 26 (4.4) | 32 (8.1) |
| Death or transplant | 241 (75.8) | 366 (64.8) | 283 (47.3) | 234 (58.9) |
| Median survival months (IQR) | 39.3 (13.4-70.6) | 53.2 (24.8-92.5) | 40.6 (19.9-75.5) | 48 (15-105) |

\* n for missing data: FVC (n=6); DLCO (n=31); GAP Stage (n=6)
\*\* n for missing data: FVC (n=241); DLCO (n=264); GAP Stage (n=241)
\*\*\* n for missing data: FVC (n=30); DLCO (n=41); GAP Stage (n=30)
\*\*\*\* n for missing data: FVC (n=4); DLCO (n=9); GAP stage (n=4)

### Genome-wide survival analysis

After filtering, 7,873,835 variants in the US dataset, 8,591,398 variants in the UK dataset, and 8,620,496 variants in the UUS dataset were tested for TFS association. Quintile-quintile plots for each stage I cohort suggested acceptable inflation (**Figure E7)**. After stratifying stage I cohorts by minor allele frequency (MAF), inflation was higher for rare variants compared to low and high frequency variants, but within an acceptable range for each group (λ<1.1) (**Figure E8**). For meta-analysis, 9,075,629 variants were tested for TFS association in the aggregated stage I cohorts. One hundred and sixty-one independent SNPs had Wald *p*<0.05 in at least two datasets with the same direction of effect and p<5.0×10^−5^ in stage I meta-analysis (**Figure 1**). Of those, 158 satisfied the proportional hazards assumption and advanced for stage II testing (**Table E1**).

**Figure 1.**
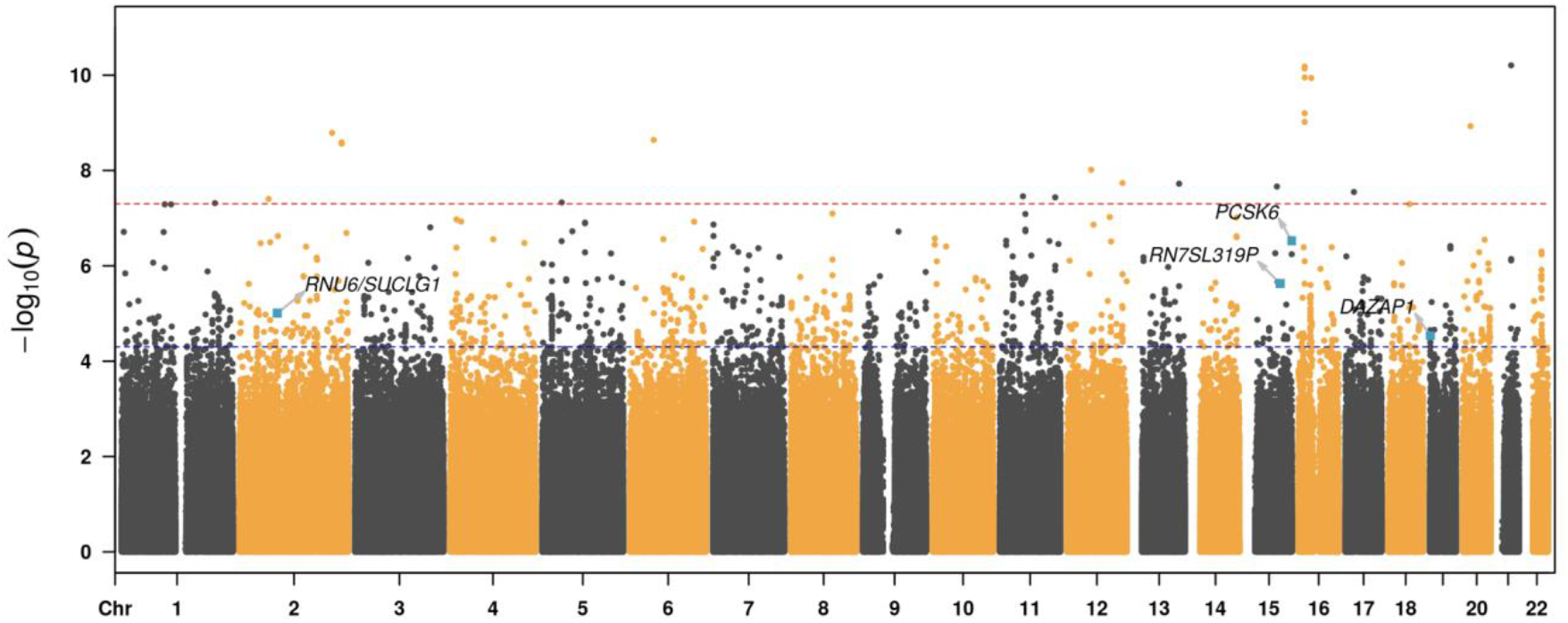
Manhattan plot of stage I gene variants associated with IPF survival. SNPs reaching nominal significance (*p*<5×10^−5^) fall above the blue line and those reaching genome-wide significance (*p*<5.0×10^−8^) fall above the red line.

Genotype data was available in the Vanderbilt University cohort for 154 of the 158 variants advanced from stage I. Six variants were associated with TFS in the Vanderbilt University cohort at *p*<0.05, including four with consistent effect direction that strengthened in TFS association after meta-analysis (**Table 2**; **Table E2**). These four were rs184498750 near Succinate-CoA Ligase GDP/ADP-Forming Subunit Alpha (*SUCLG1*), rs60514164 near ubiquitin-conjugating enzyme E2Q family member 2 (*UBE2Q2*), rs35647788 in an intron of Proprotein Convertase Subtilisin/Kexin Type 6 (*PCSK6*), and rs3893252 in an intron of Deleted In Azoospermia-Associated Protein 1 (*DAZAP1*) (**Table 2**). Of these, rs35647788 (*PCSK6*) showed strong TFS association in stage I (HR 4.76; 95% CI 2.62-8.64; *p*=2.96×10^−7^) and stage II (HR 3.12; 95% CI 1.37-7.11; *p*=6.70×10^−3^) cohorts and crossed the genome-wide significance threshold in meta-analysis (HR 4.11; 95% CI 2.54-6.67; *p*=9.45×10^−9^).With the exception of rs60514164 (*UBE2Q2*) (MAF=8%), these SNPs were low frequency, with MAF of ∼1% in the study population. Regional association plots for each of the four variants are shown in **Figure E9**.

**Table 2.** Results for the SNPs nominally associated across stages and with same direction of effects on IPF survival.

|  |  |  |  |  |  |  |  |  |  |  |  |  |  |  |
| --- | --- | --- | --- | --- | --- | --- | --- | --- | --- | --- | --- | --- | --- | --- |
| Variant location (hg19) and nearest gene |  |  |  |  |  | Stage I results |  |  |  | Stage II results |  |  | Meta-analysis |  |
| Chr | Position | SNP rsID | Gene | REF | EA | R<sup>2</sup> | EAF | HR [95% CI] | P-value | EAF | HR [95% CI] | P-value | HR [95% CI] | P-value |
| 2 | 84291167 | rs184498750 | *SUCLG1* | G | T | 0.86 | 1.1% | 3.11<br>[1.88-5.15] | 9.83x10<sup>-6</sup> | 1.2% | 2.05<br>[1.79-4.18] | 0.049 | 2.71<br>[1.79-4.08] | 2.07x10<sup>-6</sup> |
| 15 | 76081200 | rs60514164 | *UBE2Q2* | C | T | 0.77 | 7.0% | 1.60<br>[1.32-1.95] | 2.35x10<sup>-6</sup> | 7.7% | 1.42<br>[1.04-1.93] | 0.026 | 1.55<br>[1.31-1.83] | 2.23x10<sup>-7</sup> |
| 15 | 101914234 | rs35647788 | *PCSK6* | C | T | 0.93 | 0.8% | 4.76<br>[2.62-8.64] | 2.96x10<sup>-7</sup> | 0.8% | 3.12<br>[1.37-7.11] | 6.72x10<sup>-3</sup> | 4.11<br>[2.54-6.67] | 9.45x10<sup>-9</sup> |
| 19 | 1412985 | rs3893252 | *DAZAP1* | C | T | 0.82 | 0.6% | 3.57<br>[1.97-6.49] | 2.91x10<sup>-5</sup> | 1.5% | 2.09<br>[1.05-4.15] | 0.036 | 2.84<br>[1.81-4.45] | 5.81x10<sup>-6</sup> |

Abbreviations: Ref = reference allele; EA = effect allele; R<sup>2</sup>= lowest imputation quality value across studies; EAF = effect allele frequency; HR = hazard ratio; CI = confidence interval

In multivariable analysis, each variant except rs3893252 (*DAZAP1*) maintained survival association after adjustment for relevant confounders of IPF survival (**Table E3**). Among patients with the rs35647788 (*PCSK6*) variant, all were heterozygotes (**Table E4**) and were evenly distributed across centers comprising the UK and UUS cohorts. No rs35647788 (*PCSK6*) variants were observed in the US cohort despite good imputation quality (r^2^=0.74). In sensitivity analysis of the *PCSK6* variant, results were consistent when censoring transplants (**Table E5**). *In silico* testing revealed functional effects for each of the four variants associated with TFS after stage II meta-analysis **(Table E6)**. None of the variants identified had known association with fibrotic lung disease.

### PCSK6 tissue expression

Morphologic assessment of histological sections from lung tissue in patients with IPF were compared with control subjects without fibrotic lung disease. In IPF lung, cytoplasmic PCSK6 expression localized to ciliated epithelial cells and alveolar epithelial cells and was markedly higher than PCSK6 expression in non-IPF control sections (**Figure 2**). Western Blot confirmed the presence of only a single PCSK6 band (**Figure E10**). Relative staining intensity was two-fold higher in IPF lung samples (n=86) compared with non-IPF controls (n=9) (p<0.001) (**Figure 2**).

**Figure 2.**
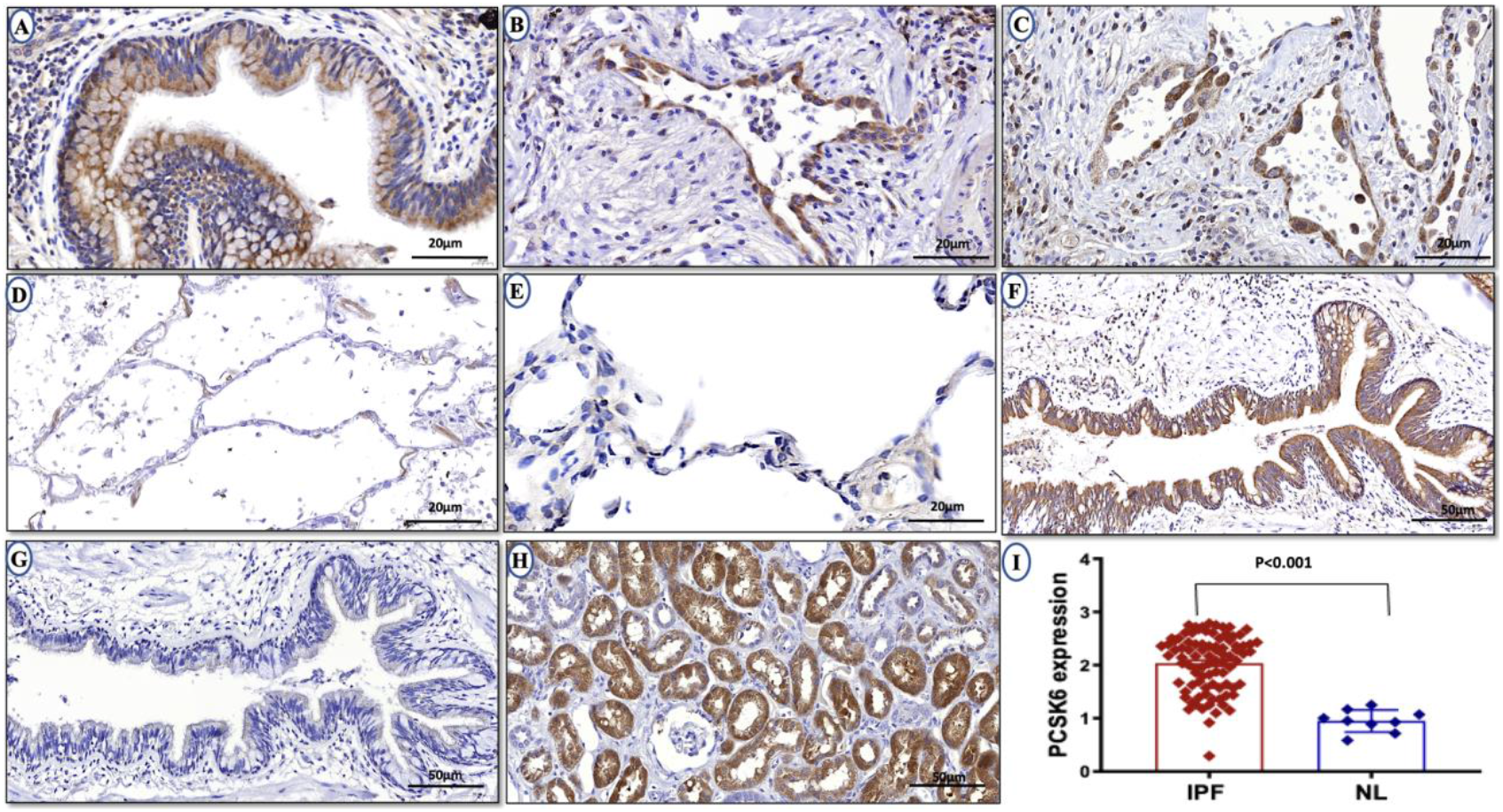
PCSK6 immunohistochemistry showed increased cytoplasmic PCSK6 expression in ciliated epithelial cells (A) and alveolar epithelial cells (B-C) compared with normal lung control sections (D-E). Parallel IPF section sections (F-G) confirm increased PCSK6 staining (F) when compared to control section (G). Human kidney positive control (H) is provided for reference. Comparison of relative PCSK6 staining intensity (I) demonstrated significantly higher mean intensity in IPF lungs when compared to non-IPF control lungs (*p*<0.001). Magnification is 20um magnification for panels A-E and 50um for panels F-G.

### PCSK6 clinical outcome association

Increased PCSK6 protein staining score was associated with reduced TFS in those with available survival data (n=71), with staining scores above the median associated with greater than 2-fold increased risk of death or lung transplant (HR 2.41; 95% CI 1.12-5.16; p=0.024) (**Figure 3a**). When assessing *PCSK6* gene expression in the COMET (n=75), Imperial College (n=55) and University of Chicago (n=45) cohorts, increasing PCSK6 expression was associated with increased mortality risk in each cohort, with each one-unit increased associated with greater than three-fold increased risk of death or lung transplant in meta-analysis (HR 3.43; 95% CI 1.62-7.25; p=0.0012) (**Figure 3b**). When assessing PCSK6 plasma concentration in patients with IPF from UC-Davis (n=138) and UChicago (n=181), increasing plasma PCSK6 concentration was associated with reduced TFS, with each one-unit change in log-transformed plasma concentration associated with a nearly 50% increase in outcome risk (HR 1.47; 95% CI 1.14, 1.89; p=0.0031). These results were consistent across UC-Davis (HR 1.47, 95% CI 1.14-1.89; p=0.08) and UChicago (HR 1.34, 95% CI 0.92-1.94; p=0.12) cohorts. After stratification of the combined cohort by tertiles, those with PCSK6 concentration in the highest tertile displayed significantly worse survival than those in the second and third tertiles (*p*=0.0018) (**Figure 3c**).

**Figure 3.**
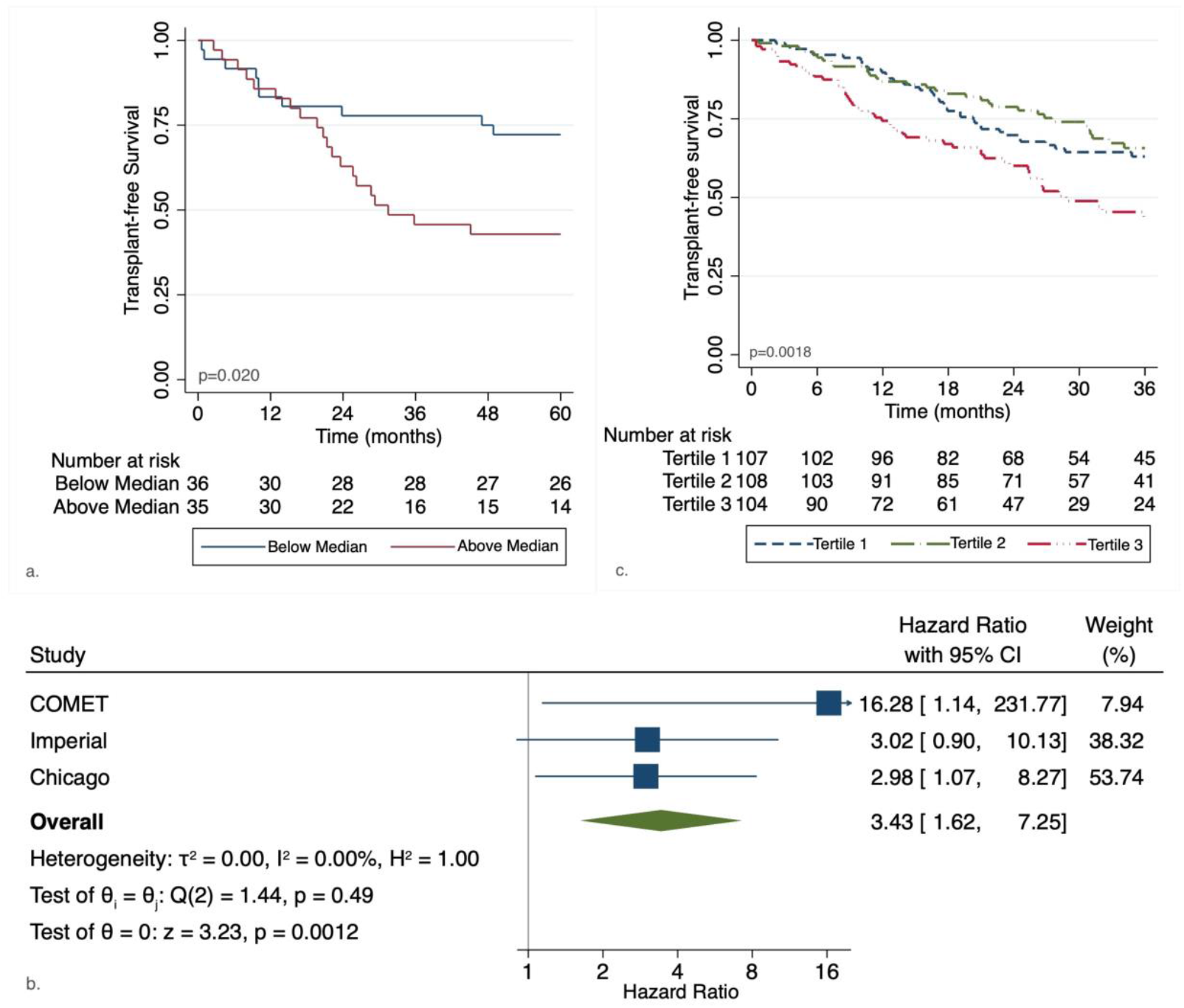
Relationship between PCSK6 and clinically relevant IPF endpoints. Higher PCSK6 staining intensity (a), peripheral blood gene expression (b) and circulating plasma protein concentration (c) are associated with reduced transplant-free survival.

### Overlap between IPF risk and transplant free survival

Variants previously associated with IPF risk^6-10^ were investigated for outcome association (**Supplementary Methods**). None of the 15 genetic variants with previously associated with IPF risk^6-10^ were associated with TFS after Bonferroni correction (*p*=0.0033) (**Table E7**). As previously reported,^16,22^ individuals with the *MUC5B* promoter polymorphism (rs3570590) displayed better overall survival, though this did not reach significance after adjustment for multiple testing. None of the four validated survival variants were associated with differential IPF risk (**Table E8**). When combing the effect of thousands of IPF risk variants in a polygenic risk score, this risk score was not significantly associated with TFS for any significance threshold used (**Figure E11**), again suggesting that variants that affect disease risk may have little impact on survival times after diagnosis.

## Discussion

In this investigation, we conducted the first GWAS of IPF survival, identifying a variant intronic to *PCSK6* that associated with differential TFS at genome-wide significance in two independent IPF cohorts totaling nearly two thousand patients. We subsequently found that PCSK6 protein was highly expressed in IPF lung tissue, localizing to alveolar epithelial cells, which play a key role in IPF onset and progression.^23^ Finally, we found that PCSK6 lung staining, peripheral blood gene expression and circulating plasma concentration negatively correlated with TFS across independent IPF cohorts. To our knowledge, this study is the first to systematically identify gene variants associated with IPF survival and the first to identify PCSK6 as a potential therapeutic target in IPF.

*PCSK6*, also called *PACE4*, encodes a widely expressed calcium-dependent serine endoprotease with known expression in alveoli and alveolar macrophages.^24^ PCSK6 is a critical mediator of TGF-*β* processing and is crucial for reproduction, embryological development and blood pressure regulation.^25-29^ A *PCSK6* gene variant has been implicated in the development of hypertension^29^ and dysregulated *PCSK6* gene expression has been linked to vascular disease,^30-32^ and cardiac remodeling following myocardial ischemia.^33^ These cardiovascular remodeling effects make PCSK6 of particular relevance to IPF, as *PCSK6* overexpression can lead to increased collagen I and III deposition, TGF-*β* activation and extracellular matrix formation,^33,34^ which are cardinal features of IPF pathogenesis.^2^ Additionally, PCSK6 may bind tissue inhibitors of metalloproteinases (TIMPs),^35^ potentially counteracting pro-fibrotic metalloproteinases.^36^

PCSK6 dysregulation has also been implicated in the development of cancer of the lung,^37^ breast,^38^ ovary^39^ thyroid^40^ and prostate.^41^ PCSK6 has been shown to regulate apoptosis in prostate cancer^42^ and pancreatic cancer^43^ and also linked to increased cancer cell invasiveness by enhancing bioactivity of matrix metalloproteinases and cytokines.^44^ Accordingly, PCSK6 has been proposed as an anti-tumor therapeutic target^45,46^ and a bioavailable formulation of an anti-PCSK6 molecule is currently under investigation.^47^ *In vitro* PCSK6 inhibition has already been shown to reduce fibroblast proliferation, migration and invasion in rheumatoid arthritis-associated synovitis,^48^ suggesting it as a candidate target for the treatment of pulmonary fibrosis.

The *PCSK6* variant identified in this study is rare and unlikely to singularly explain subsequent gene expression and protein findings. *In silico* analyses identified several nearby regulatory elements which may include common functional variants, with smaller effects, in linkage disequilibrium with this *PCSK6* variant. While beyond the scope of this investigation, further research is needed to explore other rare variants in *PCSK6* that may have an additive effect with the one identified, along with regulatory elements, expression quantitative trait loci and more complex structural variants that may be contributing to our findings.

This study also highlights important differences between genomic determinants of IPF susceptibility and survival, suggesting that genes involved in disease onset may be independent of those driving disease progression. No survival-associated variant identified in this analysis was associated with IPF risk. While replicating prior IPF risk association for the *MUC5B* promoter polymorphism, we found only weak association with improved survival, an observation that may be influenced by index event bias.^22^ As none of the survival-associated variants showed an association with disease risk, it is unlikely these survival results are affected by index event bias. These findings have potential implications for drug development, as genes associated with IPF survival may represent more effective therapeutic targets than those associated with IPF onset.

This study has several limitations. First, we acknowledge the relatively small sample size used to conduct this analysis. Despite being the largest genomic IPF outcome analysis reported to date, the modest size of this cohort limited our ability to identify higher frequency SNPs with modest effect sizes. Given sample size constraints for this rare disease, we pursued a two-stage approach with meta-analysis of candidate variants rather than a discovery/replication approach, which would have required substantially higher sample sizes in each cohort. The consistent effect association across cohorts and genome-wide significance for the *PCSK6* variant after meta-analysis increases confidence that this represents a true association, as does the downstream clinical outcome analysis showing PCSK6 gene expression and protein concentration to be associated with differential TFS. Next, there were likely differences between cohorts used to conduct this study. Survival was best among patients in the UUS cohort, which may reflect selection bias, as these patients had less follow-up compared with those in the US and UK datasets. Additionally, a large proportion of patients comprising the UUS dataset were recruited after the US approval of pirfenidone and nintedanib, which might explain the better overall survival and patients recruited before 2012 may have been exposed to potentially harmful immunosuppression.^49^ The timing of study recruitment relative to IPF diagnosis was also unknown, which could have influenced results. Finally, we only assessed individuals of European ancestry. Validation of results is required in patients of other ancestral backgrounds.

## Conclusion

Here we present results from the first GWAS of IPF survival conducted to date. This study sheds important light on the genetics of IPF progression and identified novel variants which may contribute to this process, including rs35647788 in an intron of *PCSK6*.Downstream analysis demonstrated PCSK6 protein lung staining, peripheral blood gene expression and circulating plasma concentration to be associated with reduced IPF survival, suggesting PCSK6 may serve as a potential therapeutic target in patients with IPF.

## Data Availability

GWAS Summary statistics for this study can be accessed from https://github.com/genomicsITER/PFgenetics and will be made available at GWAS Catalog

## Acknowledgements

We thank all the patients that contributed biologic samples and clinical data for this research.

