## Supplemental Methods for "PCSK6 and Survival in Idiopathic Pulmonary Fibrosis"

**PCSK6 and Genomic Determinants of Idiopathic Pulmonary Fibrosis Survival**

### Online data supplement

**Supplemental Methods** …………………… 2-4

### Supplemental Methods

#### Functional effects of survival-associated variants

*In silico* assessments were used to infer the biological effect of variants associated with TFS after stage II testing. Gene prioritization was based on Open Targets Genetics scores. Potential regulatory effects were then assessed for epigenetic mechanisms [HaploReg, RegulomeDB], long-distance genomic interactions [Capture Hi-C Plotter (CHiCP), considering a default score of 5 to select interactions], tissue-specific cis eQTLs and DNase I sensitivity QTLs (dsQTLs) [GTEx, considering a tissue-specific $P \leq 0.05$ as threshold, and SNPDelScore]. Effect of survival associated variants on other traits was assessed using PhenoScanner ($P<0.001$).

#### Survival association for IPF susceptibility-associated variants

To assess the association between IPF survival and variants previously linked to IPF-risk,[1-4] polygenic risk score (PRS) analysis was performed using PRSice[5] (v1.25) to determine genetic overlap between IPF risk and survival. An IPF susceptibility PRS was calculated using weights from a previous IPF risk GWAS[1] and tested for its association with TFS in individuals in the UUS study who were not included in the IPF risk GWAS. Independent variants (selected through LD clumping with $r^2 \leq 0.1$) associated with IPF risk at $P \leq 0.001$[5] comprised the initial PRS with threshold adjustments made to identify the PRS that explained the highest proportion of IPF risk. The final PRS was then tested for TFS association with the aforementioned Cox proportional hazards regression model using the survival package in R v3.5.3. Given its large effect on IPF risk, analyses were then repeated excluding variants near the *MUC5B* promoter polymorphism (within 500 kb of rs35705950).

#### PCSK6 Immunohistochemistry

The tissue samples were obtained after informed consent and local ethics approval (South East Scotland SAHSC Bioresource-reference number 06/S1101/41; Brompton Node samples- reference number 15/SC/0101; Papworth Node Samples reference number 08/H0304/56+5; non-diseased controls-reference number (Q)GM030404 and Nottingham BRC samples- reference number 08/H0407/1). Formalin-Fixed Paraffin Embedded (FFPE) tissue samples were cut, 5µm thick, on positively charged Leica Surgipath X-tra slides. IHC staining was performed using the Novocastra Novolink™ Polymer Detection Systems kit (RE7280-K, Leica, Biosystems, Newcastle, UK) as previously described.[6] In brief, tissue sections were deparaffinised with xylene and rehydrated through 100% ethanol. Heat-induced (pH=6) citrate antigen retrieval was performed and rabbit PCSK6 antibody (HPA004774, Atlas antibodies, Sweden; 1:200 dilution of stock antibody) was incubated overnight at 4°C. 3-3′ Diaminobenzidine tetrahydrochloride (Novolink DAB substrate buffer plus) was used as the chromogen. Slides were counterstained with Novolink haematoxylin for 6 min, dehydrated and cover slipped. Normal kidney tissue was used as a positive tissue control, whereas no primary antibody was used as a negative control.

**PCSK6 Western blotting**

Cell Lysis Buffer (Cell Signalling, USA) supplemented with protease inhibitor cocktail (Sigma, USA) was used to collect the cell protein and the western blotting protein concentrations were determined by BCA assay using a commercially available kit (Thermo Fisher Scientific, Waltham, MA), according to the manufacturer's instructions. 20 µg protein were loaded per lane of a 4-12%, pre-cast Bis-Tris gradient gels (Thermo Fisher Scientific, Waltham, MA), subject to electrophoresis, and transferred onto a nitrocellulose membrane (Merck, GE10600002). Membranes were blocked for 1 h in 5% non-fat milk in tris-buffered saline containing 0.1% Tween, pH=7.4 (TBST). Membranes were incubated with rabbit anti-PCSK6 (HPA004774, Atlas antibodies, Sweden–1:1000 dilution of stock antibody) diluted in the blocking buffer for overnight at 4°C. A loading control of mouse Anti-β-Actin was also used to demonstrate protein loading (Merck; A5441; Mouse monoclonal- Anti-β-Actin antibody at 1:100000 dilution of stock antibody). Following day membranes was washed in TBST, incubated with an anti-mouse-HRP and anti-rabbit-HRP conjugated secondary antibodies (Dako, USA) at 1:2500 for 1hr at room temperature. Visualization was performed with Clarity Max™ ECL Substrate (Biorad, UK) on a Licor C-DiGit.

**PCSK6 Plasma Testing**

Stored frozen plasma in ethylenediaminetetraacetic acid aliquots from UC-Davis (n=187) and UChicago (n=139) were thawed and processed at UC-Davis in institutional batches. PCSK6 concentration was determined by enzyme-linked immunosorbent assay (ELISA) using a commercially available kit purchased from MyBioSource (San Diego, CA). The kit was run according to the manufacturer's instructions. Briefly, standards, undiluted plasma samples and a horseradish peroxidase-conjugated detection antibody were added to 96-well plates pre-coated with capture antibody before incubation for 1 h at 37°C. The wells were then washed and developed with chromogen solution included with the kit and immediately read at 450 nM using an ELISA plate reader. Data are reported as ng/mL. Intra- and inter-assay variability was controlled for using control standards on each plate. Protein concentration was log transformed for outcome modeling.

**PCSK6 Protein Staining Intensity**

The immunohistochemically stained slides were scanned using a ScanScope XT Slide Scanner (LeicaAperio Technologies, Vista, CA, USA) under 20x objective magnification (0.5 µm resolution) using Pannoramic Viewer (3DHISTECH Ltd Budapest, Hungary) slide viewing software. Both the percentage of staining and staining intensity of PCSK6 expression in lung sections were individually assessed. For PCSK6 quantification, the following scoring system of seven high-power fields at X40 per tissue section were used:

- **Score 0:** No cells stained
- **Score 0.5**: 1-10 cells stained at low intensity
- **Score 1.0:** 1-10 cells stained at high intensity
- **Score 1.5:** 11-25 cells stained at low intensity
- **Score 2.0:** 11-25 cells stained at high intensity
- **Score 2.5:** ≥26 cells stained at low intensity

**- Score 3.0:** ≥26 cells stained at high intensity

To normalize for varying numbers of regions of interest per slide, the mean score per slide was calculated. A Mann-Whitney U-test was used to compare between IPF and control lung samples. Statistical analysis was performed using GraphPad Prism 7.0 (GraphPad Software, San Diego, CA, USA).

**PCSK6 Plasma Testing**

Stored frozen plasma in ethylenediaminetetraacetic acid aliquots from UC-Davis (n=187) and UChicago (n=139) were thawed and processed at UC-Davis in institutional batches. PCSK6 concentration was determined by enzyme-linked immunosorbent assay (ELISA) using a commercially available kit purchased from MyBioSource (San Diego, CA). The kit was run according to the manufacturer's instructions. Briefly, standards, undiluted plasma samples and a horseradish peroxidase-conjugated detection antibody were added to 96-well plates pre-coated with capture antibody before incubation for 1 h at 37°C. The wells were then washed and developed with chromogen solution included with the kit and immediately read at 450 nM using an ELISA plate reader. Data are reported as ng/mL. Intra- and inter-assay variability was controlled for using control standards on each plate. Protein concentration was log transformed for outcome modeling.

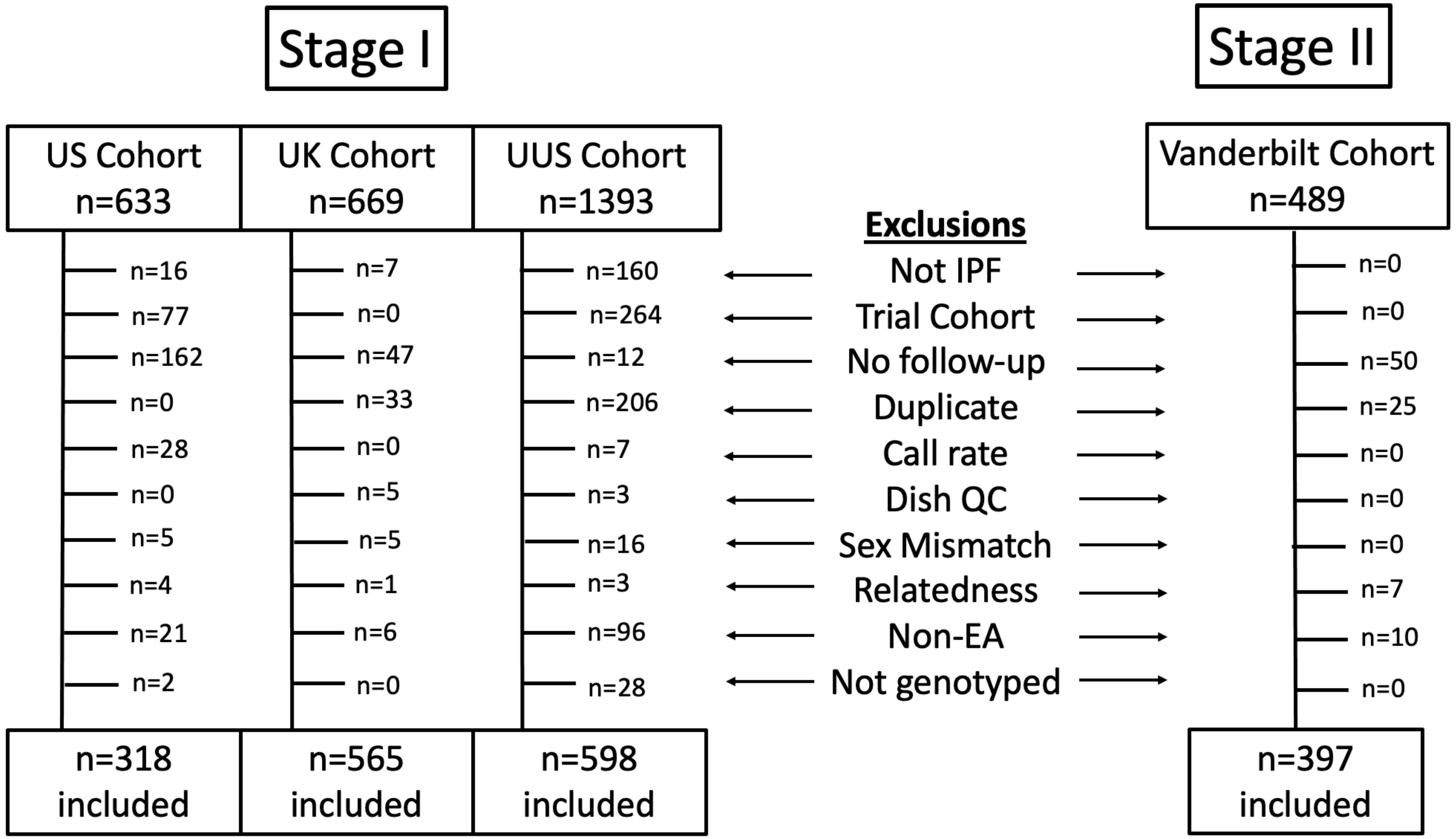

**Figure E1**. Quality control and filtering results

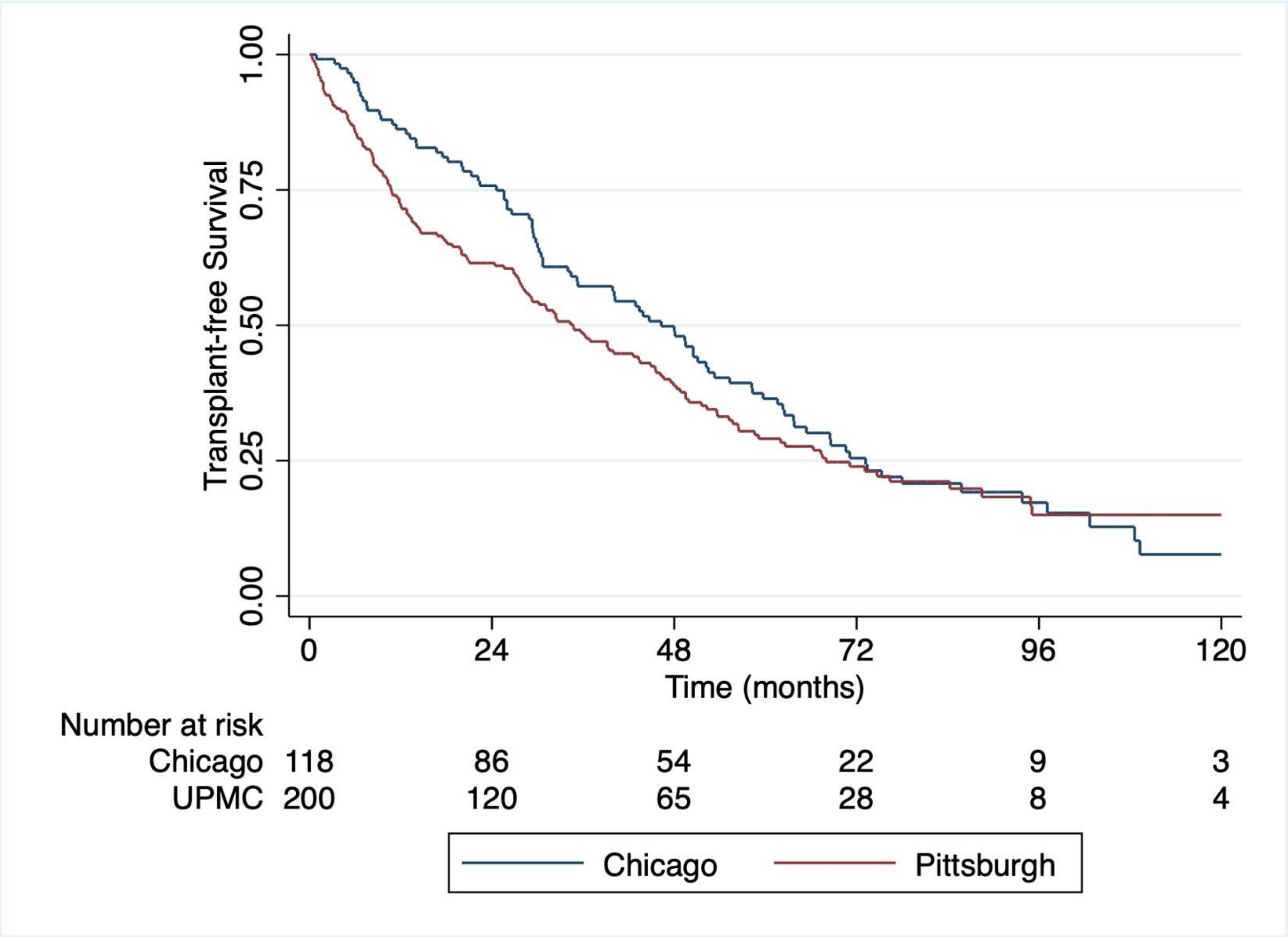

**Figure E2.** Kaplan-Meier survival plot for IPF cohorts within US dataset.

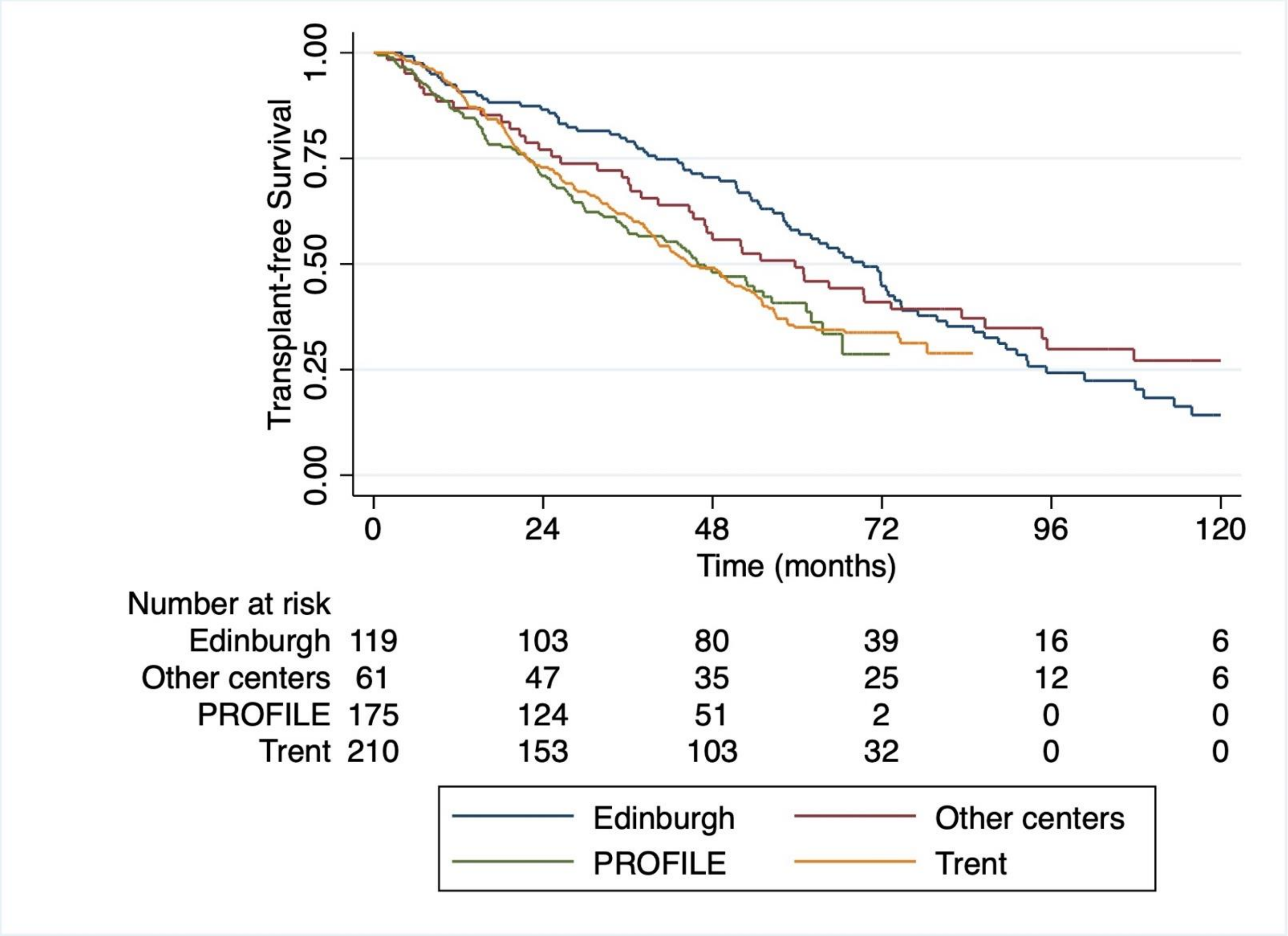

**Figure E3**. Kaplan-Meier survival plot for IPF cohorts within UK dataset.

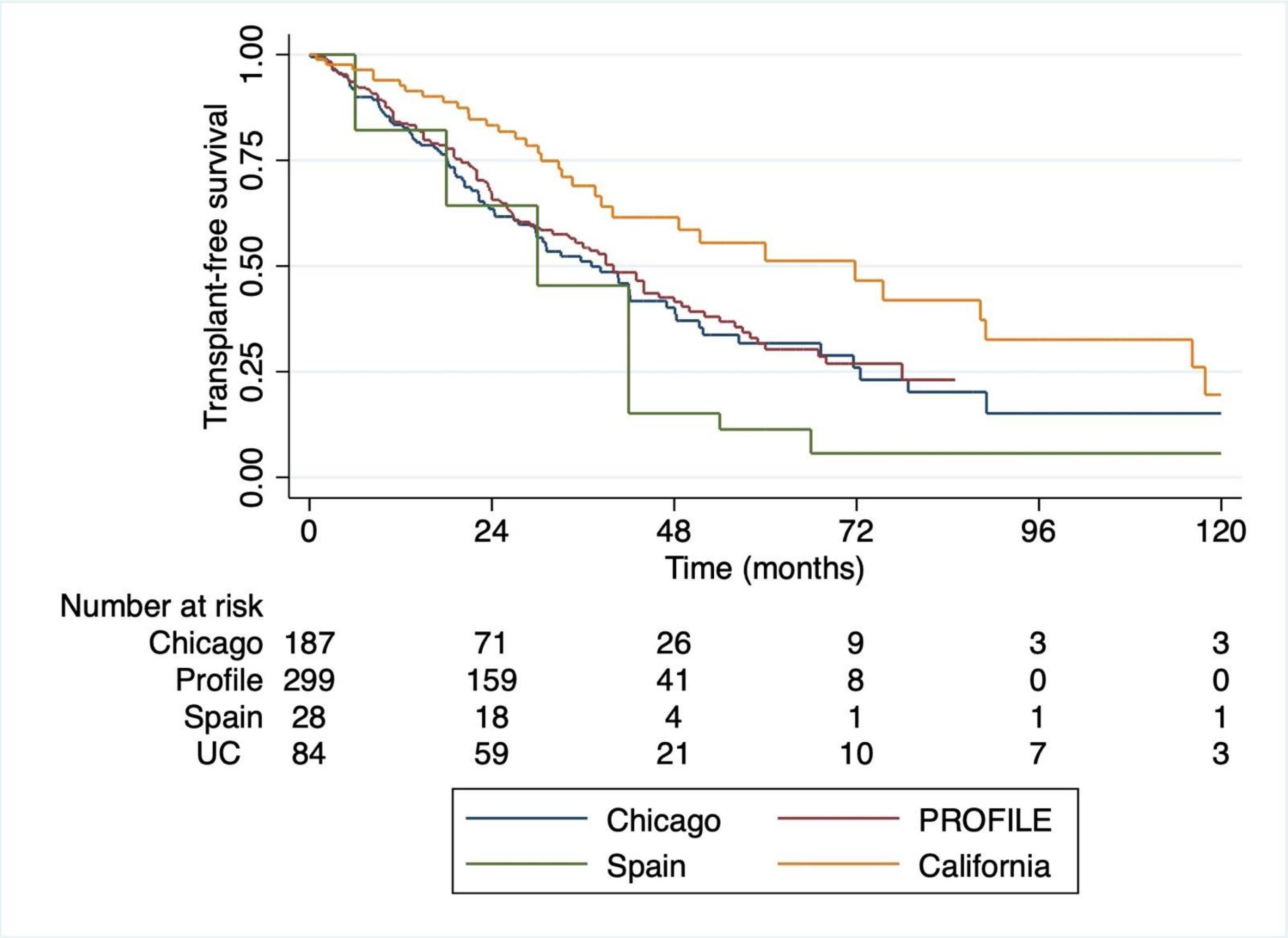

**Figure E4**. Kaplan-Meier survival plot for IPF cohorts within UUS dataset (US, UK, Spain).

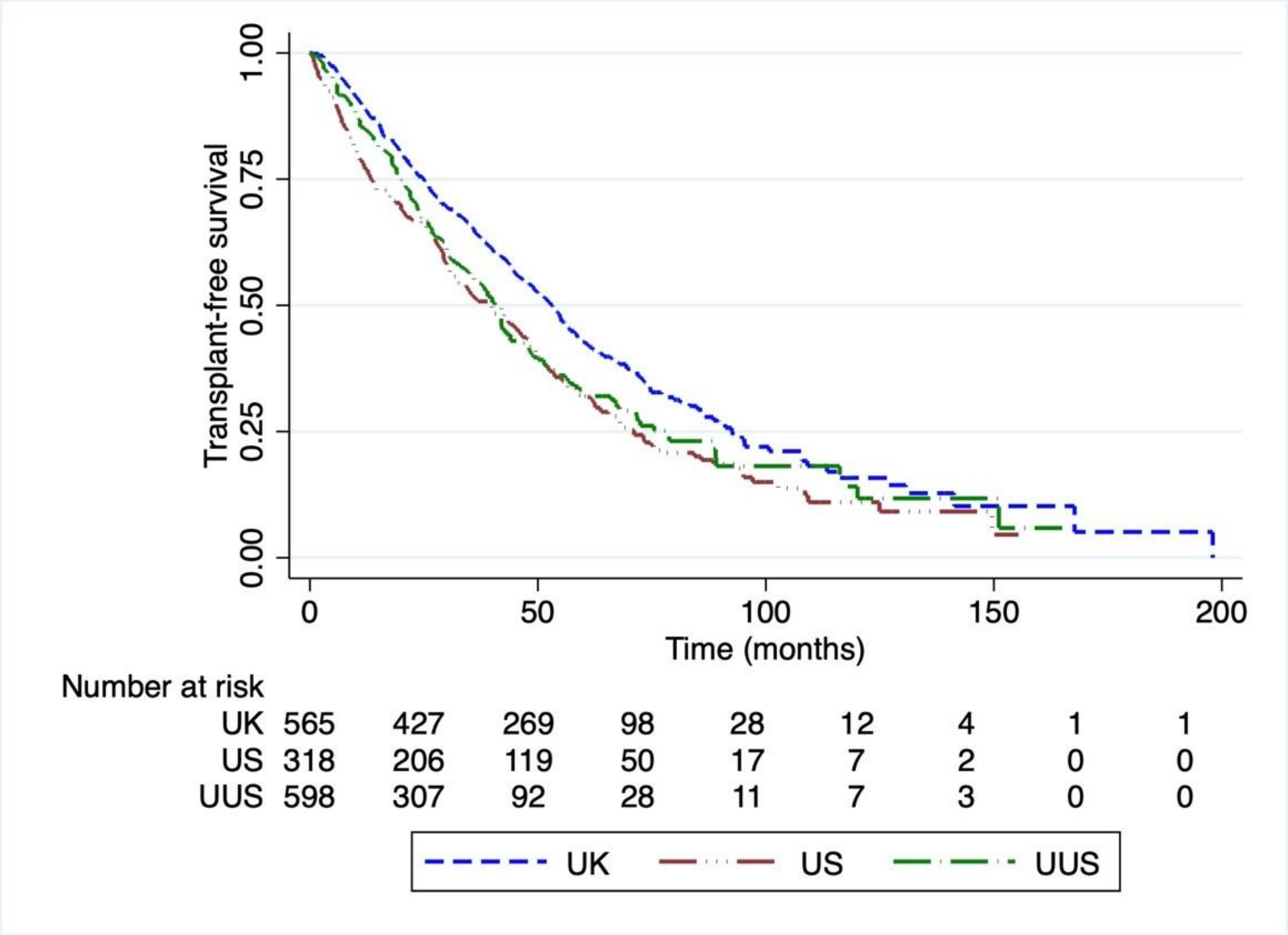

**Figure E5**. Kaplan-Meier survival plot for US, UK and UUS patients included in GWAS stage I.

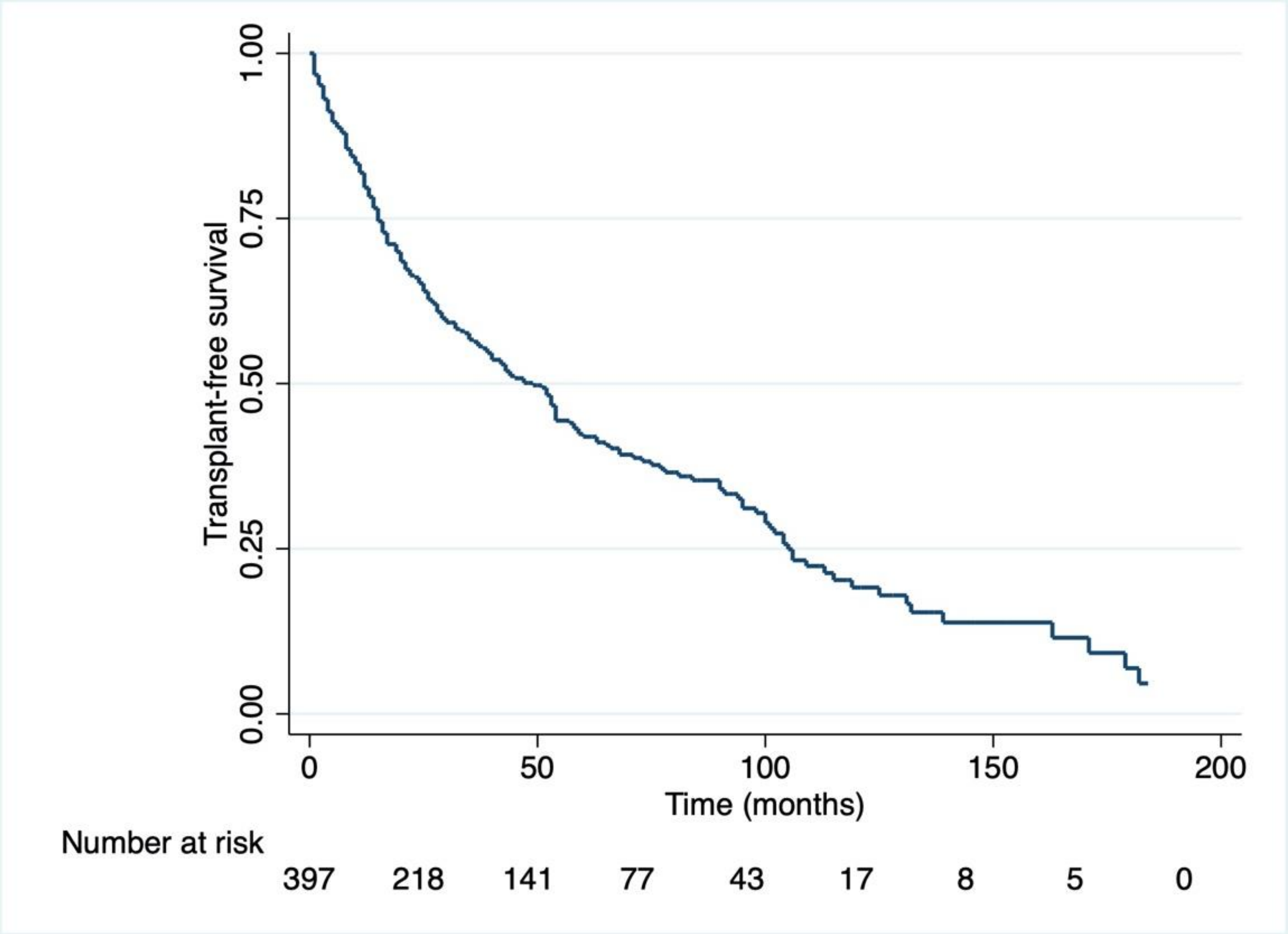

**Figure E6**. Kaplan-Meier survival plot for patients included in GWAS stage II.

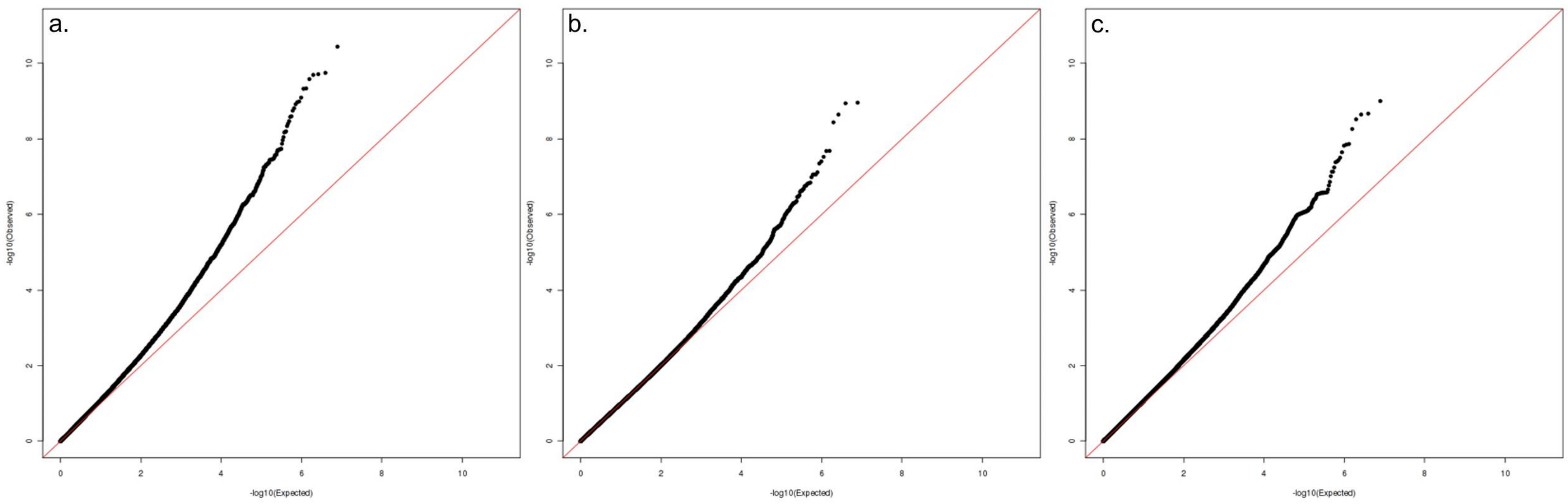

**Figure E7.** QQ plots for US ($\lambda$=1.014) (a), UK ($\lambda$=0.996) (b) and UUS ($\lambda$=1.082) (c) cohorts.

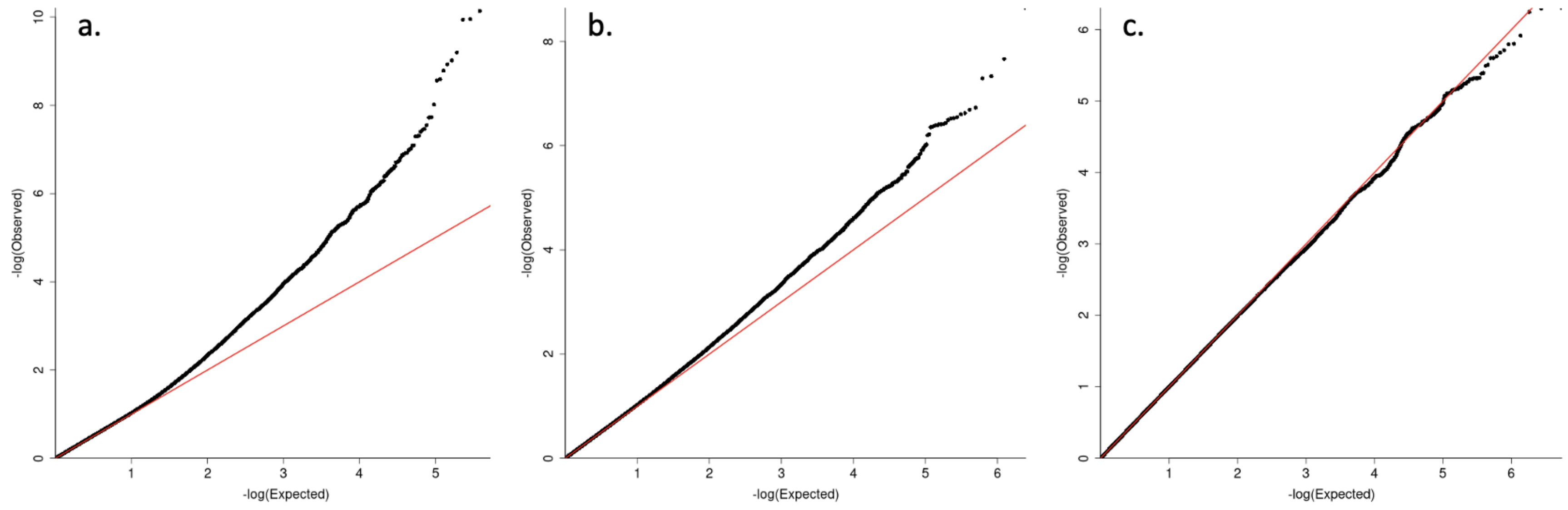

**Figure E8.** QQ plots for minor allele frequency (MAF) bins, including MAF<1% (a) (λ=1.05), MAF 1-5% (b) (λ=1.02) and MAF >5% (c) (λ=1.00).

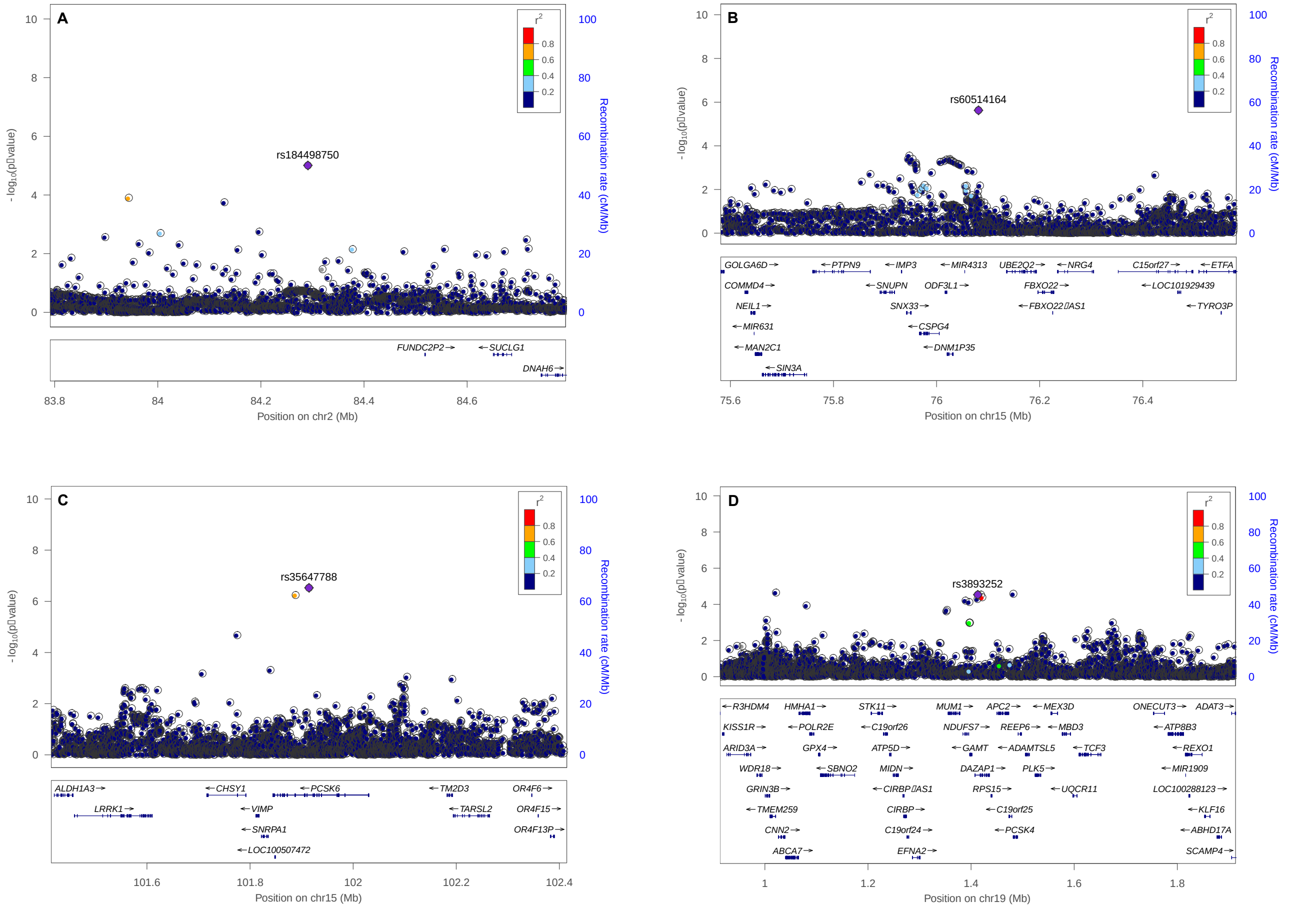

**Figure E9.** Regional plots for top variants after stage II meta-analysis. Y-axis displays -log10 transformed p-value and x-axis shows the hg19 genomic position. Estimated recombination rates (light blue line) are plotted on the right y-axis. The results for the remaining SNPs are color coded to reflect their degree of linkage disequilibrium with the leading SNP (indicated) based on pairwise $r^2$ values in Europeans.

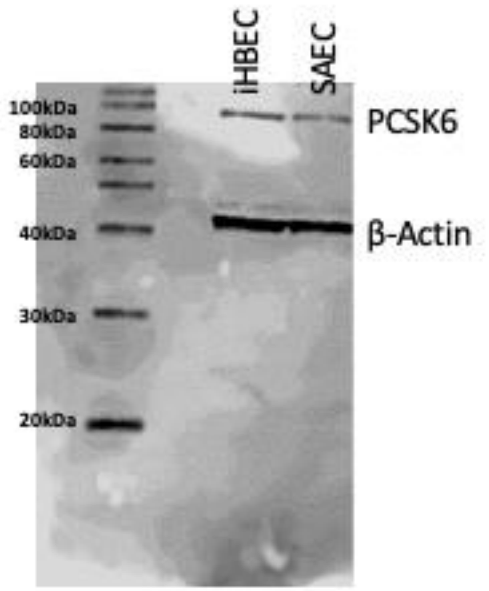

**Figure E10.** PCSK6 Western Blot. Single bands for PCSK6 (HPA004774) in the immortalized human bronchial epithelial cells (iHBECs) and small airway epithelial cells (SAEC) cells, at molecular weight of 100kDa, β-Actin (A544) was used as a loading control around 42kDa.

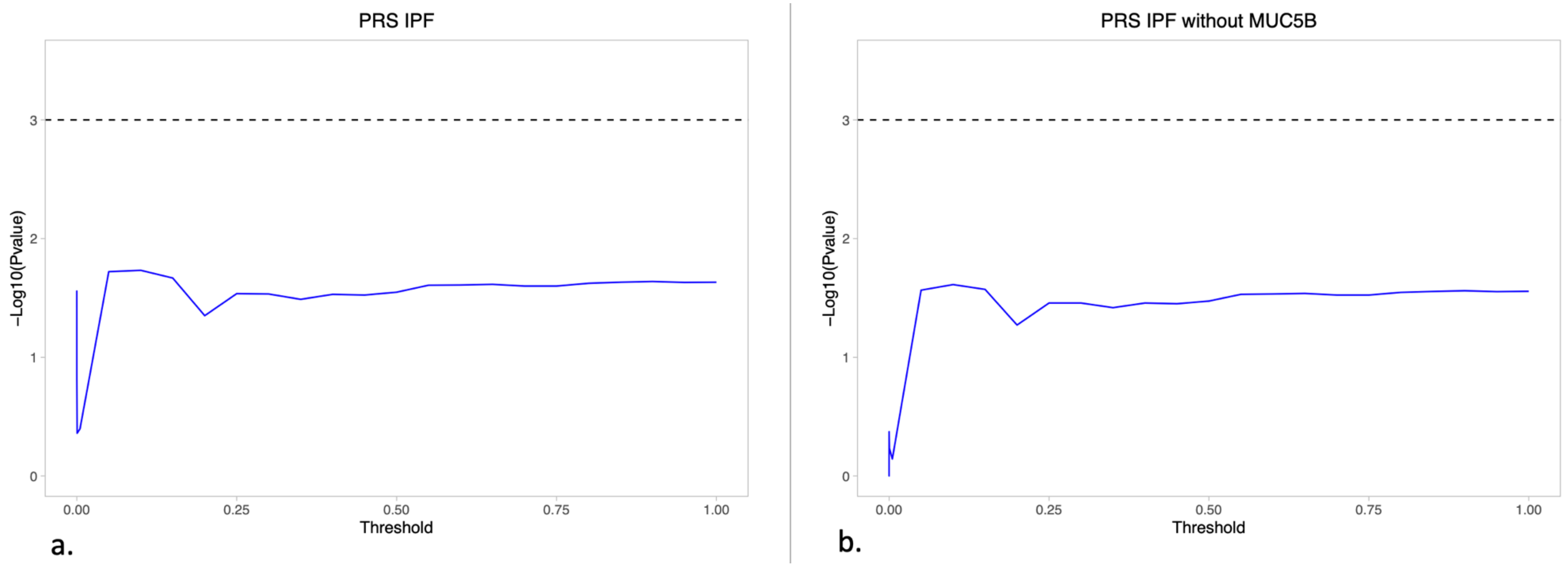

**Figure E11.** Strength of association between IPF risk polygenic risk score and transplant-free survival. The x axis shows the *p*-threshold used for determining which variants to include in the risk PRS calculation (i.e. as the value increases then more variants are included in the calculation) and the y axis shows the association of the PRS with TFS. The dotted line shows the significance threshold of $p$=0.001. Figure a) shows the association of the PRS when calculated using variants from across the genome, b) shows the strength of association when excluding the *MUC5B* region from the PRS calculation.

**Table E1**. Stage I variants of nominal significance identified in the 3-way meta-analysis

| Variant Location and Alleles | | | | | US cohort | | | | UK cohort | | | | UUS cohort | | | | Stage-I meta-analysis | |
|---|---|---|---|---|---|---|---|---|---|---|---|---|---|---|---|---|---|---|
| **Chr** | **Pos** | **SNP rsID** | **REF** | **EA** | **R2** | **EAF** | **HR [95%CI]** | **P** | **R2** | **EAF** | **HR [95%CI]** | **P** | **R2** | **EAF** | **HR [95%CI]** | **P** | **HR [95%CI]** | **P** |
| 1 | 10762349 | rs7514663 | C | T | NA | NA | NA | NA | 0.91 | 0.0141 | 2.11 [1.16-3.84] | 1.39E-2 | 0.92 | 0.0107 | 4.10 [2.01-8.37] | 1.07E-4 | 2.74 [1.72-4.36] | 2.15E-5 |
| 1 | 94606086 | rs185926659 | G | T | 0.92 | 0.0050 | 6.68 [1.87-23.9] | 3.44E-3 | 0.93 | 0.0072 | 5.24 [2.51-11.0] | 1.05E-5 | NA | NA | NA | NA | 5.55 [2.91-10.6] | 1.95E-7 |
| 1 | 100493151 | rs148192333 | T | C | NA | NA | NA | NA | 0.93 | 0.0111 | 2.61 [1.36-5.02] | 4.07E-3 | 0.89 | 0.0075 | 3.81 [1.63-8.92] | 2.08E-3 | 2.98 [1.76-5.05] | 4.75E-5 |
| 1 | 170154993 | rs111749519 | A | G | 0.81 | 0.0124 | 2.7 [1.15-6.36] | 2.30E-2 | 0.92 | 0.0094 | 2.91 [1.43-5.93] | 3.31E-3 | 0.89 | 0.0080 | 2.68 [1.14-6.3] | 2.32E-2 | 2.79 [1.74-4.47] | 2.17E-5 |
| 1 | 188069617 | rs190285902 | C | G | NA | NA | NA | NA | 0.87 | 0.0137 | 1.91 [1.03-3.56] | 4.05E-2 | 0.85 | 0.0084 | 6.93 [2.95-16.3] | 8.72E-6 | 2.92 [1.76-4.86] | 3.69E-5 |
| 1 | 188151083 | rs145551873 | G | A | 0.62 | 0.0117 | 4.72 [1.79-12.4] | 1.66E-3 | 0.95 | 0.0226 | 1.88 [1.15-3.06] | 1.15E-2 | 0.91 | 0.0125 | 2.75 [1.33-5.69] | 6.43E-3 | 2.35 [1.61-3.45] | 1.05E-5 |
| 1 | 200768521 | rs11589092 | G | A | 0.97 | 0.0975 | 1.50 [1.08-2.07] | 1.43E-2 | 0.98 | 0.0851 | 1.38 [1.07-1.77] | 1.19E-2 | 0.97 | 0.0736 | 1.45 [1.06-1.99] | 2.03E-2 | 1.43 [1.20-1.70] | 4.68E-5 |
| 1 | 200861595 | rs968549 | C | T | 0.95 | 0.9014 | 0.65 [0.47-0.89] | 8.45E-3 | 1.00 | 0.9138 | 0.72 [0.56-0.92] | 1.01E-2 | 1.00 | 0.9284 | 0.71 [0.52-0.98] | 3.52E-2 | 0.7 [0.59-0.83] | 4.29E-5 |
| 1 | 203327373 | rs115684501 | C | T | NA | NA | NA | NA | 0.92 | 0.0082 | 3.08 [1.4-6.77] | 5.01E-3 | 0.89 | 0.0059 | 6.94 [2.22-21.7] | 8.55E-4 | 3.95 [2.05-7.61] | 3.97E-5 |
| 1 | 214247966 | rs115702239 | C | A | NA | NA | NA | NA | 0.86 | 0.0082 | 4.07 [1.71-9.68] | 1.49E-3 | 0.83 | 0.0092 | 3.97 [1.82-8.67] | 5.51E-4 | 4.02 [2.22-7.27] | 4.36E-6 |
| 1 | 222319103 | rs112891824 | C | T | 0.64 | 0.0100 | 4.97 [1.44-17.2] | 1.11E-2 | 0.99 | 0.0159 | 2.26 [1.26-4.06] | 6.53E-3 | 1.00 | 0.0159 | 2.19 [1.20-4.01] | 1.04E-2 | 2.41 [1.60-3.62] | 2.25E-5 |
| 1 | 242551121 | rs34473966 | A | C | NA | NA | NA | NA | 0.89 | 0.0116 | 3.48 [1.85-6.55] | 1.11E-4 | 0.88 | 0.0115 | 2.42 [1.12-5.22] | 2.49E-2 | 3.03 [1.84-4.97] | 1.24E-5 |
| 2 | 38114198 | rs114698213 | C | T | NA | NA | NA | NA | 0.91 | 0.0126 | 2.97 [1.61-5.48] | 4.79E-4 | 0.78 | 0.0146 | 2.36 [1.21-4.61] | 1.17E-2 | 2.69 [1.70-4.26] | 2.43E-5 |
| 2 | 38249347 | rs150622385 | G | A | NA | NA | NA | NA | 0.85 | 0.0146 | 2.30 [1.25-4.25] | 7.71E-3 | 0.80 | 0.0116 | 3.88 [1.94-7.73] | 1.21E-4 | 2.87 [1.80-4.57] | 9.72E-6 |
| 2 | 43602198 | rs185306441 | C | A | NA | NA | NA | NA | 0.93 | 0.0137 | 2.01 [1.05-3.84] | 3.51E-2 | 0.90 | 0.0120 | 4.41 [2.26-8.58] | 1.28E-5 | 2.90 [1.80-4.65] | 1.07E-5 |
| 2 | 47162883 | rs115376562 | T | C | NA | NA | NA | NA | 0.88 | 0.0063 | 2.81 [1.24-6.38] | 1.32E-2 | 0.85 | 0.0053 | 12.41 [4.81-32.0] | 1.88E-7 | 5.15 [2.75-9.67] | 3.34E-7 |
| 2 | 54643373 | rs150765591 | T | G | NA | NA | NA | NA | 0.78 | 0.0106 | 2.87 [1.41-5.86] | 3.73E-3 | 0.79 | 0.0105 | 4.21 [1.78-9.98] | 1.09E-3 | 3.33 [1.9-5.82] | 2.43E-5 |
| 2 | 84291167 | rs184498750 | G | T | NA | NA | NA | NA | 0.86 | 0.0131 | 3.23 [1.76-5.92] | 1.49E-4 | 0.86 | 0.0079 | 2.86 [1.20-6.81] | 1.75E-2 | 3.11 [1.88-5.15] | 9.83E-6 |
| 2 | 138337791 | rs149029025 | G | A | NA | NA | NA | NA | 0.93 | 0.0069 | 3.82 [1.68-8.70] | 1.41E-3 | 0.91 | 0.0055 | 4.04 [1.54-10.6] | 4.45E-3 | 3.91 [2.07-7.38] | 2.65E-5 |
| 2 | 142907569 | rs142306180 | C | A | 0.92 | 0.0102 | 10.55 [4.16-26.8] | 7.11E-7 | NA | NA | NA | NA | 0.95 | 0.0053 | 2.89 [1.03-8.16] | 4.43E-2 | 5.88 [2.85-12.2] | 1.63E-6 |
| 2 | 172941078 | rs2357323 | T | C | 0.86 | 0.0053 | 4.51 [1.1-18.6] | 3.68E-2 | NA | NA | NA | NA | 0.84 | 0.0089 | 4.57 [2.09-10.0] | 1.44E-4 | 4.56 [2.23-9.32] | 3.23E-5 |
| 2 | 172942149 | rs56923699 | G | A | 0.86 | 0.0053 | 4.84 [1.17-20.0] | 2.93E-2 | NA | NA | NA | NA | 0.85 | 0.0092 | 4.55 [2.09-9.93] | 1.38E-4 | 4.62 [2.26-9.42] | 2.59E-5 |
| 2 | 173111445 | rs182271851 | A | G | 0.81 | 0.0085 | 3.08 [1.19-7.98] | 2.09E-2 | NA | NA | NA | NA | 0.94 | 0.0051 | 8.84 [3.69-21.2] | 1.05E-6 | 5.48 [2.79-10.8] | 7.49E-7 |
| 2 | 226721440 | rs377036225 | G | A | NA | NA | NA | NA | 0.89 | 0.0105 | 5.25 [2.45-11.3] | 2.04E-5 | 0.87 | 0.0157 | 2.00 [1.06-3.78] | 3.23E-2 | 3.03 [1.84-4.99] | 1.39E-5 |
| 2 | 226758606 | rs142266733 | A | G | NA | NA | NA | NA | 0.85 | 0.0118 | 4.16 [2.06-8.41] | 6.95E-5 | 0.81 | 0.0135 | 2.35 [1.15-4.81] | 1.95E-2 | 3.18 [1.91-5.31] | 9.25E-6 |
| 2 | 240384770 | rs188643417 | G | A | NA | NA | NA | NA | 0.83 | 0.0085 | 2.42 [1.07-5.49] | 3.43E-2 | 0.88 | 0.0111 | 3.81 [2.02-7.18] | 3.49E-5 | 3.19 [1.91-5.33] | 9.73E-6 |

**Table E1**. Stage I variants of nominal significance identified in the 3-way meta-analysis

| Variant Location and Alleles | | | | | US cohort | | | | UK cohort | | | | UUS cohort | | | | Stage-I meta-analysis | |
|---|---|---|---|---|---|---|---|---|---|---|---|---|---|---|---|---|---|---|
| Chr | Pos | SNP rsID | REF | EA | R2 | EAF | HR [95%CI] | P | R2 | EAF | HR [95%CI] | P | R2 | EAF | HR [95%CI] | P | HR [95%CI] | P |
| 2 | 241253363 | rs116377902 | G | C | NA | NA | NA | NA | 0.95 | 0.0070 | 3.82 [1.76-8.29] | 6.75E-4 | 0.94 | 0.0071 | 2.74 [1.19-6.29] | 1.73E-2 | 3.30 [1.85-5.87] | 4.96E-5 |
| 2 | 241397273 | rs75406448 | A | G | 0.67 | 0.0301 | 1.99 [1.04-3.81] | 3.79E-2 | 0.97 | 0.0379 | 1.79 [1.22-2.62] | 2.72E-3 | 0.97 | 0.0300 | 1.72 [1.10-2.69] | 1.80E-2 | 1.80 [1.37-2.35] | 2.29E-5 |
| 3 | 20681518 | rs73177699 | T | G | 0.89 | 0.0174 | 2.16 [1.11-4.2] | 2.30E-2 | 0.92 | 0.0122 | 2.46 [1.29-4.69] | 6.07E-3 | 0.92 | 0.0068 | 4.56 [1.90-10.9] | 6.75E-4 | 2.68 [1.76-4.07] | 4.27E-6 |
| 3 | 59911730 | rs77895452 | C | A | 0.82 | 0.0059 | 6.68 [2.13-21.0] | 1.13E-3 | NA | NA | NA | NA | 0.91 | 0.0050 | 4.49 [1.55-13.0] | 5.52E-3 | 5.39 [2.39-12.10] | 4.91E-5 |
| 3 | 69741636 | rs139644937 | T | A | 0.96 | 0.0062 | 7.63 [2.33-25.0] | 7.84E-4 | 1.00 | 0.0071 | 2.74 [1.31-5.74] | 7.68E-3 | 0.99 | 0.0071 | 2.59 [1.04-6.43] | 4.08E-2 | 3.23 [1.91-5.48] | 1.34E-5 |
| 3 | 75034442 | rs142255892 | C | T | NA | NA | NA | NA | 0.78 | 0.0107 | 4.23 [2.07-8.64] | 7.39E-5 | 0.71 | 0.0108 | 2.88 [1.30-6.36] | 9.13E-3 | 3.59 [2.09-6.16] | 3.54E-6 |
| 3 | 79693632 | rs36022026 | A | C | 0.95 | 0.0616 | 1.83 [1.25-2.66] | 1.76E-3 | 0.97 | 0.0675 | 1.47 [1.09-1.98] | 1.19E-2 | 0.97 | 0.0722 | 1.4 [1.01-1.94] | 4.30E-2 | 1.52 [1.25-1.85] | 2.36E-5 |
| 3 | 112828302 | rs2178398 | C | G | 0.99 | 0.1511 | 0.73 [0.56-0.95] | 2.13E-2 | 0.96 | 0.1884 | 0.74 [0.60-0.91] | 4.97E-3 | 0.97 | 0.1836 | 0.75 [0.59-0.94] | 1.48E-2 | 0.74 [0.64-0.85] | 2.02E-5 |
| 3 | 145344805 | rs368666349 | C | A | NA | NA | NA | NA | 0.91 | 0.0069 | 2.25 [1.22-4.17] | 9.74E-3 | 0.90 | 0.0096 | 4.04 [2.02-8.09] | 7.91E-5 | 2.88 [1.80-4.60] | 9.57E-6 |
| 4 | 11752169 | rs191600377 | T | C | NA | NA | NA | NA | 0.81 | 0.0050 | 6.99 [2.34-20.9] | 5.02E-4 | 0.85 | 0.0078 | 3.59 [1.65-7.79] | 1.26E-3 | 4.54 [2.37-8.69] | 5.15E-6 |
| 4 | 11982650 | rs17260001 | G | A | NA | NA | NA | NA | 0.90 | 0.0056 | 8.38 [3.24-21.70] | 1.18E-5 | 0.90 | 0.0073 | 3.22 [1.30-7.95] | 1.13E-2 | 5.17 [2.65-10.10] | 1.49E-6 |
| 4 | 12091599 | rs192085162 | G | A | NA | NA | NA | NA | 0.90 | 0.0056 | 5.38 [2.21-13.05] | 2.02E-4 | 0.92 | 0.0065 | 3.29 [1.26-8.59] | 1.53E-2 | 4.33 [2.23-8.41] | 1.51E-5 |
| 4 | 66572997 | rs142199841 | T | C | 0.86 | 0.0084 | 3.62 [1.36-9.62] | 9.86E-3 | 0.92 | 0.0054 | 4.87 [2.05-11.60] | 3.35E-4 | NA | NA | NA | NA | 4.31 [2.22-8.34] | 1.52E-5 |
| 4 | 74459567 | rs79914686 | T | C | NA | NA | NA | NA | 0.82 | 0.0054 | 6.24 [2.24-17.40] | 4.70E-4 | 0.81 | 0.0055 | 3.58 [1.24-10.3] | 1.86E-2 | 4.82 [2.27-10.20] | 4.12E-5 |
| 4 | 76495474 | rs28641522 | A | G | 0.83 | 0.0067 | 11.56 [3.45-38.7] | 7.14E-5 | NA | NA | NA | NA | 0.86 | 0.0061 | 3.17 [1.26-7.98] | 1.44E-2 | 5.08 [2.36-10.9] | 3.22E-5 |
| 4 | 122351715 | rs6850444 | G | C | 0.90 | 0.3205 | 0.79 [0.64-0.97] | 2.61E-2 | 0.99 | 0.3401 | 0.81 [0.69-0.96] | 1.65E-2 | 0.98 | 0.3709 | 0.74 [0.62-0.89] | 1.26E-3 | 0.78 [0.7-0.87] | 1.14E-5 |
| 4 | 166645872 | rs71618464 | T | C | NA | NA | NA | NA | 0.93 | 0.0100 | 3.19 [1.56-6.50] | 1.46E-3 | 0.91 | 0.0074 | 4.62 [1.90-11.2] | 7.11E-4 | 3.66 [2.08-6.44] | 6.56E-6 |
| 4 | 175369612 | rs139698405 | C | T | NA | NA | NA | NA | 0.81 | 0.0079 | 4.03 [1.57-10.40] | 3.69E-3 | 0.81 | 0.0059 | 4.40 [1.76-11.00] | 1.58E-3 | 4.21 [2.15-8.24] | 2.69E-5 |
| 4 | 175401693 | rs190563090 | A | G | NA | NA | NA | NA | 0.84 | 0.0093 | 2.77 [1.16-6.63] | 2.23E-2 | 0.83 | 0.0053 | 11.41 [4.27-30.50] | 1.20E-6 | 5.03 [2.59-9.78] | 1.88E-6 |
| 5 | 36581565 | rs138830243 | C | T | 0.82 | 0.0109 | 2.93 [1.1-7.81] | 3.20E-2 | 0.86 | 0.0116 | 3.07 [1.57-5.98] | 9.88E-4 | 0.84 | 0.0120 | 2.41 [1.10-5.27] | 2.73E-2 | 2.81 [1.77-4.46] | 1.11E-5 |
| 5 | 96796936 | rs138577491 | T | C | NA | NA | NA | NA | 0.86 | 0.0063 | 5.85 [2.44-14.0] | 7.56E-5 | 0.88 | 0.0061 | 3.95 [1.49-10.5] | 5.92E-3 | 4.95 [2.55-9.62] | 2.30E-6 |
| 5 | 106109149 | rs571176768 | G | C | 0.92 | 0.0079 | 2.63 [1.04-6.63] | 4.07E-2 | 0.97 | 0.0098 | 3.17 [1.62-6.22] | 7.63E-4 | 0.95 | 0.0081 | 2.86 [1.27-6.44] | 1.14E-2 | 2.95 [1.85-4.68] | 4.77E-6 |
| 5 | 151623819 | rs191657347 | A | G | 0.92 | 0.0069 | 3.74 [1.37-10.2] | 1.02E-2 | 0.93 | 0.0054 | 4.51 [1.91-10.70] | 6.06E-4 | NA | NA | NA | NA | 4.19 [2.15-8.16] | 2.63E-5 |
| 5 | 154587673 | rs536809954 | G | A | NA | NA | NA | NA | 0.89 | 0.0055 | 5.62 [2.38-13.30] | 8.48E-5 | 0.86 | 0.0075 | 3.73 [1.54-9.06] | 3.62E-3 | 4.65 [2.48-8.72] | 1.75E-6 |
| 5 | 154820302 | rs13172001 | G | A | NA | NA | NA | NA | 0.87 | 0.0120 | 3.35 [1.77-6.36] | 2.16E-4 | 0.87 | 0.0094 | 2.37 [1.12-5.02] | 2.38E-2 | 2.92 [1.78-4.79] | 2.26E-5 |
| 6 | 26118992 | rs116594669 | C | G | 0.76 | 0.0107 | 3.52 [1.36-9.09] | 9.33E-3 | 0.89 | 0.0080 | 3.57 [1.53-8.37] | 3.35E-3 | 0.89 | 0.0143 | 2.02 [1.00-4.06] | 4.90E-2 | 2.77 [1.71-4.49] | 3.71E-5 |

**Table E1**. Stage I variants of nominal significance identified in the 3-way meta-analysis

| Variant Location and Alleles | | | | | US cohort | | | | UK cohort | | | | UUS cohort | | | | Stage-I meta-analysis | |
|---|---|---|---|---|---|---|---|---|---|---|---|---|---|---|---|---|---|---|
| Chr | Pos | SNP rsID | REF | EA | R2 | EAF | HR [95%CI] | P | R2 | EAF | HR [95%CI] | P | R2 | EAF | HR [95%CI] | P | HR [95%CI] | P |
| 6 | 75389935 | rs145442342 | G | A | 0.74 | 0.0135 | 2.69 [1.18-6.11] | 1.84E-2 | 0.93 | 0.0102 | 2.43 [1.20-4.93] | 1.38E-2 | 0.92 | 0.0062 | 4.27 [1.66-11.0] | 2.67E-3 | 2.86 [1.77-4.62] | 1.65E-5 |
| 6 | 77044422 | rs140590158 | C | T | NA | NA | NA | NA | 0.92 | 0.0206 | 2.61 [1.59-4.29] | 1.53E-4 | 0.90 | 0.0190 | 2.2 [1.27-3.81] | 4.87E-3 | 2.43 [1.67-3.53] | 3.62E-6 |
| 6 | 84945776 | rs116863455 | G | A | NA | NA | NA | NA | 0.90 | 0.0086 | 4.38 [2.17-8.85] | 3.73E-5 | 0.86 | 0.0075 | 3.13 [1.20-8.15] | 1.98E-2 | 3.92 [2.21-6.96] | 3.16E-6 |
| 6 | 94466663 | rs149057325 | A | T | 0.93 | 0.0288 | 1.81 [1.08-3.02] | 2.35E-2 | 0.98 | 0.0262 | 2.00 [1.25-3.21] | 3.86E-3 | 0.98 | 0.0289 | 1.62 [1.03-2.55] | 3.58E-2 | 1.80 [1.36-2.39] | 4.42E-5 |
| 6 | 94498243 | rs143246710 | C | T | 0.94 | 0.0177 | 2.23 [1.18-4.24] | 1.39E-2 | 0.97 | 0.0161 | 2.09 [1.19-3.65] | 9.84E-3 | 0.97 | 0.0153 | 2.11 [1.17-3.81] | 1.27E-2 | 2.14 [1.50-3.04] | 2.38E-5 |
| 6 | 94531687 | rs144768626 | A | G | 0.93 | 0.0176 | 2.3 [1.2-4.39] | 1.17E-2 | 0.97 | 0.0168 | 2.08 [1.21-3.59] | 8.30E-3 | 0.96 | 0.0159 | 1.96 [1.08-3.55] | 2.63E-2 | 2.10 [1.48-2.97] | 3.43E-5 |
| 6 | 102917669 | rs139568075 | C | T | NA | NA | NA | NA | 0.80 | 0.0063 | 4.39 [1.81-10.6] | 1.07E-3 | 0.80 | 0.0084 | 3.25 [1.44-7.35] | 4.59E-3 | 3.75 [2.03-6.93] | 2.35E-5 |
| 6 | 116041139 | rs35483630 | G | A | NA | NA | NA | NA | 0.89 | 0.0060 | 4.47 [1.76-11.40] | 1.64E-3 | 0.86 | 0.0098 | 3.72 [1.87-7.40] | 1.76E-4 | 3.98 [2.26-7.03] | 1.80E-6 |
| 6 | 116377677 | rs181590625 | G | A | NA | NA | NA | NA | 0.90 | 0.0052 | 3.20 [1.11-9.24] | 3.12E-2 | 0.88 | 0.0088 | 3.71 [1.81-7.59] | 3.29E-4 | 3.54 [1.92-6.50] | 4.89E-5 |
| 6 | 139248572 | rs535860622 | T | C | 0.83 | 0.0068 | 3 [1.03-8.72] | 4.35E-2 | NA | NA | NA | NA | 0.89 | 0.0058 | 8.84 [3.35-23.40] | 1.10E-5 | 5.44 [2.57-11.50] | 9.73E-6 |
| 6 | 143298800 | rs149730644 | C | T | 0.85 | 0.0066 | 4.49 [1.33-15.2] | 1.58E-2 | NA | NA | NA | NA | 0.95 | 0.0054 | 11.47 [4.50-29.30] | 3.26E-7 | 8.13 [3.74-17.70] | 1.19E-7 |
| 6 | 152786447 | rs62427038 | T | C | NA | NA | NA | NA | 0.82 | 0.0086 | 3.44 [1.57-7.54] | 1.97E-3 | 0.83 | 0.0117 | 2.95 [1.38-6.27] | 5.08E-3 | 3.19 [1.83-5.55] | 4.28E-5 |
| 6 | 164521835 | rs188589004 | G | A | NA | NA | NA | NA | 0.82 | 0.0071 | 2.96 [1.35-6.48] | 6.72E-3 | 0.83 | 0.0066 | 5.47 [2.13-14.10] | 4.25E-4 | 3.76 [2.03-6.94] | 2.34E-5 |
| 7 | 1065220 | rs183625998 | A | G | NA | NA | NA | NA | 0.95 | 0.0054 | 6.22 [2.60-14.90] | 3.95E-5 | 0.92 | 0.0055 | 4.54 [1.93-10.66] | 5.13E-4 | 5.33 [2.86-9.93] | 1.36E-7 |
| 7 | 1120020 | rs192803195 | C | T | NA | NA | NA | NA | 0.91 | 0.0051 | 6.83 [2.68-17.4] | 5.68E-5 | 0.90 | 0.0051 | 3.79 [1.6-9.01] | 2.50E-3 | 5.03 [2.63-9.63] | 1.04E-6 |
| 7 | 74953934 | rs868964452 | A | T | NA | NA | NA | NA | 0.62 | 0.0180 | 3.21 [1.73-5.97] | 2.19E-4 | 0.64 | 0.0125 | 2.21 [1.01-4.84] | 4.68E-2 | 2.8 [1.71-4.59] | 4.17E-5 |
| 7 | 75344084 | rs117388086 | C | A | NA | NA | NA | NA | 0.91 | 0.0110 | 2.37 [1.19-4.71] | 1.42E-2 | 0.91 | 0.0158 | 3.02 [1.67-5.46] | 2.70E-4 | 2.71 [1.71-4.29] | 2.23E-5 |
| 7 | 75358307 | rs111489307 | T | G | 0.53 | 0.0137 | 3.19 [1.23-8.27] | 1.68E-2 | 0.98 | 0.0104 | 3.33 [1.69-6.59] | 5.34E-4 | 0.97 | 0.0092 | 2.83 [1.31-6.11] | 7.96E-3 | 3.13 [1.98-4.96] | 1.20E-6 |
| 7 | 101163592 | rs149406562 | C | A | NA | NA | NA | NA | 0.93 | 0.0129 | 3.8 [2.10-6.87] | 9.88E-6 | 0.94 | 0.0098 | 2.76 [1.30-5.85] | 8.21E-3 | 3.38 [2.11-5.43] | 4.26E-7 |
| 7 | 103029848 | rs142639517 | A | G | NA | NA | NA | NA | 0.89 | 0.0082 | 4.02 [1.96-8.25] | 1.47E-4 | 0.91 | 0.0050 | 2.95 [1.25-6.95] | 1.35E-2 | 3.56 [2.03-6.23] | 8.79E-6 |
| 7 | 125546344 | rs74580469 | T | C | NA | NA | NA | NA | 0.93 | 0.0085 | 3.39 [1.61-7.13] | 1.27E-3 | 0.88 | 0.0125 | 3.11 [1.50-6.45] | 2.30E-3 | 3.25 [1.91-5.53] | 1.36E-5 |
| 7 | 148106376 | rs556475473 | T | C | NA | NA | NA | NA | 0.85 | 0.0062 | 5.33 [2.22-12.8] | 1.82E-4 | 0.86 | 0.0052 | 5.17 [2.01-13.3] | 6.39E-4 | 5.26 [2.73-10.10] | 6.51E-7 |
| 7 | 152982958 | rs73728959 | G | A | NA | NA | NA | NA | 0.99 | 0.0093 | 3.59 [1.70-7.57] | 8.10E-4 | 0.96 | 0.0104 | 3.51 [1.60-7.66] | 1.67E-3 | 3.55 [2.05-6.15] | 6.53E-6 |
| 8 | 9496015 | rs148904289 | T | A | NA | NA | NA | NA | 0.91 | 0.0053 | 3.54 [1.44-8.75] | 6.06E-3 | 0.93 | 0.0078 | 3.58 [1.60-8.01] | 1.87E-3 | 3.57 [1.93-6.59] | 4.97E-5 |
| 8 | 22381858 | rs113465261 | T | C | NA | NA | NA | NA | 0.90 | 0.0060 | 2.90 [1.14-7.41] | 2.58E-2 | 0.90 | 0.0051 | 5.20 [2.29-11.80] | 8.49E-5 | 3.99 [2.12-7.50] | 1.78E-5 |
| 8 | 23454598 | rs193064386 | G | T | NA | NA | NA | NA | 0.87 | 0.0067 | 3.80 [1.69-8.55] | 1.28E-3 | 0.88 | 0.0062 | 3.70 [1.53-8.92] | 3.63E-3 | 3.75 [2.04-6.89] | 2.01E-5 |

**Table E1**. Stage I variants of nominal significance identified in the 3-way meta-analysis

| Variant Location and Alleles | | | | | US cohort | | | | UK cohort | | | | UUS cohort | | | | Stage-I meta-analysis | |
|---|---|---|---|---|---|---|---|---|---|---|---|---|---|---|---|---|---|---|
| **Chr** | **Pos** | **SNP rsID** | **REF** | **EA** | **R2** | **EAF** | **HR [95%CI]** | **P** | **R2** | **EAF** | **HR [95%CI]** | **P** | **R2** | **EAF** | **HR [95%CI]** | **P** | **HR [95%CI]** | **P** |
| 8 | 23626607 | rs79562205 | C | T | NA | NA | NA | NA | 0.93 | 0.0075 | 3.64 [1.69-7.83] | 9.58E-4 | 0.91 | 0.0097 | 2.91 [1.38-6.14] | 4.93E-3 | 3.26 [1.89-5.63] | 2.16E-5 |
| 8 | 80493124 | rs116969973 | G | A | NA | NA | NA | NA | 0.94 | 0.0122 | 2.20 [1.22-3.96] | 9.08E-3 | 0.88 | 0.0126 | 4.12 [2.02-8.38] | 9.57E-5 | 2.80 [1.77-4.45] | 1.21E-5 |
| 8 | 92675281 | rs183828688 | A | G | NA | NA | NA | NA | 0.90 | 0.0072 | 7.55 [3.38-16.9] | 8.31E-7 | 0.89 | 0.0065 | 3.35 [1.34-8.39] | 9.84E-3 | 5.39 [2.91-9.98] | 8.00E-8 |
| 8 | 120287198 | rs117931584 | A | G | 0.72 | 0.0116 | 6.09 [2.43-15.3] | 1.18E-4 | NA | NA | NA | NA | 0.84 | 0.0053 | 2.95 [1.07-8.11] | 3.66E-2 | 4.37 [2.14-8.92] | 4.93E-5 |
| 9 | 4064850 | rs147994675 | C | T | 0.83 | 0.0113 | 2.47 [1.02-5.97] | 4.43E-2 | 0.89 | 0.0107 | 2.12 [1.07-4.19] | 3.15E-2 | 0.85 | 0.0130 | 3.11 [1.62-5.99] | 6.68E-4 | 2.55 [1.66-3.91] | 1.84E-5 |
| 9 | 7042171 | rs190922348 | C | T | 0.82 | 0.0161 | 2.68 [1.31-5.47] | 6.81E-3 | 0.92 | 0.0100 | 2.48 [1.26-4.88] | 8.41E-3 | 0.93 | 0.0128 | 2.26 [1.05-4.86] | 3.65E-2 | 2.48 [1.62-3.79] | 2.84E-5 |
| 9 | 14343426 | rs118165660 | G | A | NA | NA | NA | NA | 0.79 | 0.0257 | 2.53 [1.56-4.10] | 1.74E-4 | 0.76 | 0.0184 | 2.08 [1.11-3.90] | 2.26E-2 | 2.36 [1.60-3.48] | 1.55E-5 |
| 9 | 24585027 | rs113029495 | G | A | 0.87 | 0.0060 | 8.74 [2.78-27.5] | 2.04E-4 | 0.94 | 0.0062 | 3.90 [1.74-8.73] | 9.60E-4 | NA | NA | NA | NA | 5.01 [2.56-9.79] | 2.45E-6 |
| 9 | 86260066 | rs139230397 | G | C | NA | NA | NA | NA | 0.88 | 0.0056 | 4.67 [1.92-11.40] | 6.63E-4 | 0.89 | 0.0089 | 3.63 [1.55-8.47] | 2.92E-3 | 4.11 [2.20-7.70] | 9.53E-6 |
| 9 | 92489004 | rs187088797 | C | G | NA | NA | NA | NA | 0.88 | 0.0093 | 3.16 [1.59-6.29] | 1.04E-3 | 0.91 | 0.0077 | 2.89 [1.27-6.55] | 1.12E-2 | 3.05 [1.79-5.21] | 4.47E-5 |
| 10 | 4620992 | rs142562867 | G | A | 0.91 | 0.0112 | 3.48 [1.48-8.19] | 4.34E-3 | 0.97 | 0.0160 | 1.9 [1.13-3.20] | 1.55E-2 | 0.96 | 0.0142 | 2.40 [1.19-4.84] | 1.42E-2 | 2.26 [1.54-3.32] | 2.96E-5 |
| 10 | 11875431 | rs111589468 | G | A | 0.84 | 0.0058 | 7 [2.02-24.3] | 2.14E-3 | NA | NA | NA | NA | 0.89 | 0.0103 | 3.33 [1.63-6.78] | 9.42E-4 | 3.99 [2.09-7.60] | 2.60E-5 |
| 10 | 43302339 | rs139494937 | A | T | NA | NA | NA | NA | 0.93 | 0.0170 | 2.41 [1.37-4.23] | 2.19E-3 | 0.96 | 0.0086 | 3.88 [1.79-8.41] | 5.70E-4 | 2.82 [1.78-4.47] | 1.05E-5 |
| 10 | 73718436 | rs186597433 | C | T | 0.72 | 0.0110 | 3.73 [1.48-9.37] | 5.16E-3 | NA | NA | NA | NA | 0.95 | 0.0065 | 5.69 [2.45-13.20] | 5.38E-5 | 4.70 [2.45-9.02] | 3.14E-6 |
| 10 | 87849922 | rs191887805 | C | T | NA | NA | NA | NA | 0.87 | 0.0063 | 5.87 [2.44-14.10] | 7.84E-5 | 0.85 | 0.0056 | 2.66 [1.07-6.58] | 3.47E-2 | 4.06 [2.14-7.73] | 1.93E-5 |
| 10 | 120582311 | rs117054238 | T | C | NA | NA | NA | NA | 0.84 | 0.0081 | 2.33 [1.10-4.94] | 2.67E-2 | 0.81 | 0.0058 | 9.16 [3.39-24.8] | 1.25E-5 | 3.74 [2.04-6.86] | 2.08E-5 |
| 11 | 14115243 | rs78904863 | T | A | 0.87 | 0.0226 | 3.46 [1.88-6.37] | 6.56E-5 | 0.95 | 0.0197 | 1.69 [1.02-2.79] | 4.04E-2 | 0.95 | 0.0257 | 1.70 [1.07-2.69] | 2.45E-2 | 1.99 [1.47-2.70] | 9.54E-6 |
| 11 | 28663945 | rs111945608 | T | C | 0.70 | 0.0142 | 5.14 [2.34-11.3] | 4.66E-5 | 0.91 | 0.0210 | 1.64 [1.00-2.70] | 4.96E-2 | 0.93 | 0.0112 | 3.19 [1.62-6.26] | 7.56E-4 | 2.45 [1.70-3.52] | 1.43E-6 |
| 11 | 30638253 | rs189350185 | G | T | 0.95 | 0.0075 | 4.81 [1.82-12.8] | 1.58E-3 | 0.94 | 0.0073 | 3.01 [1.42-6.40] | 4.04E-3 | NA | NA | NA | NA | 3.55 [1.94-6.51] | 4.17E-5 |
| 11 | 33998481 | rs76061348 | T | C | NA | NA | NA | NA | 0.89 | 0.0124 | 2.51 [1.33-4.73] | 4.33E-3 | 0.91 | 0.0208 | 2.35 [1.37-4.04] | 1.92E-3 | 2.42 [1.59-3.69] | 3.75E-5 |
| 11 | 36108905 | rs118080683 | C | T | 0.54 | 0.0146 | 2.9 [1.1-7.64] | 3.14E-2 | 0.92 | 0.0153 | 3.01 [1.70-5.33] | 1.64E-4 | 0.89 | 0.0165 | 2.20 [1.13-4.30] | 2.11E-2 | 2.69 [1.79-4.04] | 1.78E-6 |
| 11 | 44113701 | rs186991973 | T | C | 0.87 | 0.0121 | 4.33 [1.99-9.43] | 2.24E-4 | NA | NA | NA | NA | 0.92 | 0.0064 | 2.73 [1.16-6.42] | 2.17E-2 | 3.51 [1.92-6.41] | 4.52E-5 |
| 11 | 75214357 | rs186259260 | C | T | NA | NA | NA | NA | 0.80 | 0.0053 | 3.64 [1.35-9.82] | 1.07E-2 | 0.85 | 0.0076 | 5.00 [2.24-11.20] | 8.46E-5 | 4.38 [2.31-8.31] | 5.86E-6 |
| 11 | 81937642 | rs72940008 | C | G | NA | NA | NA | NA | 0.85 | 0.0096 | 2.58 [1.31-5.11] | 6.41E-3 | 0.83 | 0.0092 | 3.64 [1.71-7.74] | 7.83E-4 | 2.99 [1.79-5.01] | 3.00E-5 |
| 11 | 123294624 | rs118091577 | G | T | NA | NA | NA | NA | 0.86 | 0.0073 | 3.24 [1.75-6.03] | 1.95E-4 | 0.86 | 0.0062 | 4.05 [1.75-9.37] | 1.10E-3 | 3.49 [2.11-5.79] | 1.22E-6 |
| 11 | 123312059 | rs559811602 | A | G | NA | NA | NA | NA | 0.92 | 0.0057 | 8.99 [3.85-21.00] | 4.04E-7 | 0.87 | 0.0051 | 3.57 [1.41-9.05] | 7.35E-3 | 6.02 [3.18-11.40] | 3.65E-8 |

**Table E1**. Stage I variants of nominal significance identified in the 3-way meta-analysis

| Variant Location and Alleles | | | | | US cohort | | | | UK cohort | | | | UUS cohort | | | | Stage-I meta-analysis | |
|---|---|---|---|---|---|---|---|---|---|---|---|---|---|---|---|---|---|---|
| Chr | Pos | SNP rsID | REF | EA | R2 | EAF | HR [95%CI] | P | R2 | EAF | HR [95%CI] | P | R2 | EAF | HR [95%CI] | P | HR [95%CI] | P |
| 11 | 126441296 | rs189162333 | A | G | 0.57 | 0.0155 | 6.04 [2.37-15.4] | 1.69E-4 | 0.67 | 0.0147 | 2.67 [1.33-5.35] | 5.61E-3 | NA | NA | NA | NA | 3.50 [1.98-6.18] | 1.51E-5 |
| 11 | 132893929 | rs192803572 | T | C | NA | NA | NA | NA | 0.86 | 0.0091 | 4.23 [2.02-8.84] | 1.29E-4 | 0.80 | 0.0061 | 3.4 [1.03-11.3] | 4.50E-2 | 4.00 [2.12-7.54] | 1.91E-5 |
| 11 | 134086356 | rs140676527 | G | A | 0.83 | 0.0062 | 4.63 [1.48-14.5] | 8.64E-3 | 0.92 | 0.0063 | 4.79 [2.06-11.20] | 2.83E-4 | NA | NA | NA | NA | 4.74 [2.37-9.46] | 1.05E-5 |
| 12 | 5522297 | rs150598029 | T | G | NA | NA | NA | NA | 0.99 | 0.0053 | 5.98 [2.49-14.40] | 6.51E-5 | 0.97 | 0.0078 | 2.36 [1.14-4.90] | 2.13E-2 | 3.52 [1.98-6.25] | 1.81E-5 |
| 12 | 6067283 | rs185814268 | A | G | NA | NA | NA | NA | 0.86 | 0.0092 | 4.66 [2.23-9.74] | 4.21E-5 | 0.85 | 0.0092 | 3.53 [1.51-8.22] | 3.51E-3 | 4.16 [2.36-7.31] | 7.79E-7 |
| 12 | 59333262 | rs4760248 | G | C | 0.99 | 0.1086 | 1.63 [1.23-2.16] | 6.93E-4 | 1.00 | 0.1090 | 1.28 [1.00-1.62] | 4.56E-2 | 0.99 | 0.0997 | 1.32 [1.01-1.72] | 3.88E-2 | 1.38 [1.18-1.61] | 4.52E-5 |
| 12 | 77326010 | rs117273906 | C | T | 0.95 | 0.0091 | 4.41 [1.8-10.8] | 1.18E-3 | 0.99 | 0.0129 | 2.21 [1.20-4.06] | 1.09E-2 | 0.99 | 0.0120 | 2.31 [1.14-4.67] | 2.06E-2 | 2.57 [1.69-3.91] | 1.07E-5 |
| 12 | 91345195 | rs61926456 | C | T | NA | NA | NA | NA | 0.89 | 0.0129 | 3.21 [1.73-5.99] | 2.35E-4 | 0.89 | 0.0103 | 2.11 [1.01-4.38] | 4.60E-2 | 2.71 [1.68-4.39] | 4.89E-5 |
| 12 | 124110116 | rs188979614 | C | T | 0.85 | 0.0100 | 3.13 [1.29-7.59] | 1.15E-2 | 0.93 | 0.0054 | 4.17 [1.77-9.8] | 1.07E-3 | 0.92 | 0.0051 | 4.77 [1.57-14.5] | 5.83E-3 | 3.88 [2.24-6.75] | 1.50E-6 |
| 12 | 124161354 | rs190737725 | C | T | 0.80 | 0.0090 | 4.16 [1.48-11.7] | 6.90E-3 | 0.94 | 0.0053 | 4.20 [1.77-9.96] | 1.12E-3 | NA | NA | NA | NA | 4.18 [2.13-8.22] | 3.32E-5 |
| 13 | 43516465 | rs192910570 | A | G | NA | NA | NA | NA | 0.82 | 0.0110 | 4.03 [2.11-7.71] | 2.57E-5 | 0.84 | 0.0106 | 2.78 [1.03-7.55] | 4.42E-2 | 3.63 [2.09-6.3] | 4.37E-6 |
| 13 | 43523271 | rs143441960 | T | C | NA | NA | NA | NA | 0.85 | 0.0144 | 2.99 [1.68-5.32] | 2.04E-4 | 0.83 | 0.0118 | 2.52 [1.06-6.01] | 3.70E-2 | 2.84 [1.75-4.63] | 2.57E-5 |
| 13 | 50380270 | rs35389309 | A | G | 0.90 | 0.0060 | 7.31 [2.39-22.4] | 4.91E-4 | NA | NA | NA | NA | 0.89 | 0.0073 | 3.69 [1.58-8.59] | 2.50E-3 | 4.72 [2.33-9.54] | 1.61E-5 |
| 13 | 77117893 | rs149658103 | C | T | 0.88 | 0.0159 | 2.23 [1.04-4.78] | 3.90E-2 | 0.96 | 0.0226 | 1.85 [1.12-3.05] | 1.58E-2 | 0.97 | 0.0144 | 2.36 [1.29-4.30] | 5.22E-3 | 2.07 [1.46-2.94] | 4.91E-5 |
| 13 | 92556834 | rs9556131 | T | A | 0.92 | 0.1107 | 1.39 [1.01-1.93] | 4.56E-2 | 0.98 | 0.0977 | 1.57 [1.24-2.00] | 2.32E-4 | 0.97 | 0.1021 | 1.33 [1.03-1.73] | 3.16E-2 | 1.45 [1.23-1.70] | 5.68E-6 |
| 14 | 53451666 | rs182533924 | A | T | NA | NA | NA | NA | 0.83 | 0.0072 | 5.29 [2.32-12.0] | 7.24E-5 | 0.81 | 0.0067 | 4.61 [1.51-14.10] | 7.21E-3 | 5.05 [2.58-9.88] | 2.25E-6 |
| 14 | 54089652 | rs187000606 | C | T | NA | NA | NA | NA | 0.88 | 0.0062 | 4.92 [2.13-11.34] | 1.84E-4 | 0.90 | 0.0062 | 3.29 [1.42-7.61] | 5.32E-3 | 4.06 [2.22-7.42] | 5.29E-6 |
| 14 | 82049712 | rs138142698 | A | G | 0.81 | 0.0087 | 3.78 [1.48-9.68] | 5.56E-3 | NA | NA | NA | NA | 0.92 | 0.0081 | 3.78 [1.74-8.22] | 7.86E-4 | 3.78 [2.02-7.07] | 3.08E-5 |
| 14 | 92700530 | rs117705014 | G | A | NA | NA | NA | NA | 0.89 | 0.0118 | 2.30 [1.26-4.22] | 6.99E-3 | 0.86 | 0.0093 | 3.98 [1.96-8.08] | 1.35E-4 | 2.87 [1.80-4.59] | 1.03E-5 |
| 15 | 76081200 | rs60514164 | C | T | 0.77 | 0.0578 | 2.11 [1.37-3.25] | 6.94E-4 | 0.92 | 0.0676 | 1.54 [1.15-2.07] | 4.22E-3 | 0.92 | 0.0848 | 1.47 [1.08-1.99] | 1.51E-2 | 1.6 [1.32-1.95] | 2.35E-6 |
| 15 | 82159556 | rs116962753 | T | C | 0.66 | 0.0102 | 3.13 [1.22-8.01] | 1.73E-2 | 0.80 | 0.0082 | 4.33 [1.99-9.40] | 2.11E-4 | NA | NA | NA | NA | 3.83 [2.08-7.04] | 1.61E-5 |
| 15 | 101914234 | rs35647788 | C | T | NA | NA | NA | NA | 0.93 | 0.0069 | 5.18 [2.35-11.42] | 4.60E-5 | 0.95 | 0.0098 | 4.25 [1.78-10.17] | 1.14E-3 | 4.76 [2.62-8.64] | 2.96E-7 |
| 16 | 13757971 | rs183941002 | G | T | NA | NA | NA | NA | 0.95 | 0.0062 | 5.21 [2.28-11.90] | 8.79E-5 | 0.92 | 0.0064 | 11.14 [4.78-26.00] | 2.34E-8 | 7.44 [4.07-13.60] | 6.60E-11 |
| 16 | 24509583 | rs28510778 | C | T | 0.75 | 0.0388 | 1.92 [1.13-3.25] | 1.57E-2 | 1.00 | 0.0517 | 1.48 [1.08-2.02] | 1.48E-2 | 1.00 | 0.0609 | 1.48 [1.10-2.00] | 9.50E-3 | 1.53 [1.25-1.88] | 4.35E-5 |
| 16 | 29437123 | rs71387661 | T | C | NA | NA | NA | NA | 0.89 | 0.0246 | 2.02 [1.30-3.16] | 1.95E-3 | 0.90 | 0.0151 | 2.63 [1.36-5.07] | 3.90E-3 | 2.19 [1.51-3.18] | 3.97E-5 |
| 16 | 29599816 | rs147189264 | G | A | NA | NA | NA | NA | 0.88 | 0.0284 | 1.92 [1.25-2.95] | 2.77E-3 | 0.88 | 0.0205 | 2.64 [1.50-4.68] | 8.27E-4 | 2.14 [1.51-3.03] | 1.67E-5 |

**Table E1**. Stage I variants of nominal significance identified in the 3-way meta-analysis

| Variant Location and Alleles | | | | | US cohort | | | | UK cohort | | | | UUS cohort | | | | Stage-I meta-analysis | |
|---|---|---|---|---|---|---|---|---|---|---|---|---|---|---|---|---|---|---|
| Chr | Pos | SNP rsID | REF | EA | R2 | EAF | HR [95%CI] | P | R2 | EAF | HR [95%CI] | P | R2 | EAF | HR [95%CI] | P | HR [95%CI] | P |
| 16 | 29603553 | rs34944462 | C | T | NA | NA | NA | NA | 0.89 | 0.0284 | 1.93 [1.26-2.95] | 2.64E-3 | 0.88 | 0.0204 | 2.65 [1.50-4.70] | 7.97E-4 | 2.15 [1.52-3.04] | 1.55E-5 |
| 16 | 57317482 | rs148521985 | C | T | NA | NA | NA | NA | 0.90 | 0.0095 | 2.75 [1.34-5.67] | 6.01E-3 | 0.89 | 0.0074 | 3.86 [1.72-8.7] | 1.10E-3 | 3.18 [1.83-5.50] | 3.70E-5 |
| 16 | 65985243 | rs144433549 | G | A | NA | NA | NA | NA | 0.88 | 0.0055 | 8.02 [3.08-20.9] | 2.08E-5 | 0.89 | 0.0064 | 3.11 [1.28-7.56] | 1.25E-2 | 4.91 [2.52-9.56] | 2.84E-6 |
| 16 | 83235650 | rs11642140 | T | C | NA | NA | NA | NA | 0.99 | 0.0584 | 1.62 [1.19-2.20] | 2.06E-3 | 0.99 | 0.0559 | 1.62 [1.17-2.26] | 4.13E-3 | 1.62 [1.29-2.04] | 3.58E-5 |
| 17 | 18860566 | rs187631060 | G | A | NA | NA | NA | NA | 0.84 | 0.0061 | 4.40 [1.78-10.9] | 1.36E-3 | 0.81 | 0.0050 | 10.27 [4.04-26.1] | 9.66E-7 | 6.53 [3.37-12.68] | 2.82E-8 |
| 17 | 32965617 | rs11650798 | G | C | 0.61 | 0.5741 | 0.69 [0.55-0.88] | 2.33E-3 | 0.98 | 0.5712 | 0.82 [0.70-0.96] | 1.40E-2 | 0.98 | 0.5763 | 0.81 [0.68-0.96] | 1.71E-2 | 0.79 [0.71-0.88] | 1.88E-5 |
| 17 | 78670545 | rs187199661 | G | A | NA | NA | NA | NA | 0.87 | 0.0061 | 3.48 [1.45-8.39] | 5.34E-3 | 0.88 | 0.0071 | 4.04 [1.75-9.33] | 1.06E-3 | 3.76 [2.02-6.97] | 2.73E-5 |
| 17 | 78806880 | rs139401580 | C | T | NA | NA | NA | NA | 0.93 | 0.0053 | 4.38 [1.82-10.6] | 9.92E-4 | 0.92 | 0.0084 | 3.37 [1.65-6.89] | 8.73E-4 | 3.76 [2.13-6.64] | 4.93E-6 |
| 18 | 12796936 | rs187303689 | G | A | NA | NA | NA | NA | 0.95 | 0.0062 | 2.63 [1.24-5.57] | 1.19E-2 | 0.93 | 0.0065 | 8.12 [3.32-19.9] | 4.56E-6 | 4.1 [2.28-7.36] | 2.32E-6 |
| 18 | 12954976 | rs191153148 | C | T | NA | NA | NA | NA | 0.88 | 0.0053 | 2.95 [1.12-7.73] | 2.79E-2 | 0.85 | 0.0055 | 9.14 [3.26-25.6] | 2.53E-5 | 4.89 [2.39-10.00] | 1.43E-5 |
| 18 | 25516518 | rs10469051 | G | C | 1.00 | 0.0739 | 1.66 [1.2-2.3] | 2.41E-3 | 1.00 | 0.0655 | 1.44 [1.09-1.90] | 9.58E-3 | 1.00 | 0.0653 | 1.38 [1.01-1.90] | 4.58E-2 | 1.48 [1.24-1.77] | 2.08E-5 |
| 18 | 30461003 | rs141194630 | T | C | 0.81 | 0.0050 | 3.41 [1.01-11.6] | 4.90E-2 | 0.87 | 0.0055 | 7.26 [3.01-17.5] | 1.00E-5 | NA | NA | NA | NA | 5.70 [2.76-11.80] | 2.58E-6 |
| 19 | 1021639 | rs150244663 | C | T | NA | NA | NA | NA | 0.88 | 0.0278 | 2.13 [1.33-3.40] | 1.54E-3 | 0.89 | 0.0210 | 2.41 [1.34-4.33] | 3.13E-3 | 2.23 [1.54-3.23] | 2.24E-5 |
| 19 | 1412576 | rs147560834 | G | A | NA | NA | NA | NA | 0.91 | 0.0103 | 2.71 [1.35-5.44] | 4.91E-3 | 0.93 | 0.0097 | 2.82 [1.45-5.50] | 2.33E-3 | 2.77 [1.69-4.52] | 4.98E-5 |
| 19 | 1412985 | rs3893252 | C | T | NA | NA | NA | NA | 0.82 | 0.0051 | 10.40 [3.60-30.0] | 1.50E-5 | 0.99 | 0.0067 | 2.18 [1.09-4.35] | 2.81E-2 | 3.57 [1.97-6.49] | 2.91E-5 |
| 19 | 1482080 | rs78359732 | C | A | NA | NA | NA | NA | 1.00 | 0.0106 | 2.52 [1.34-4.72] | 3.92E-3 | 0.99 | 0.0100 | 2.88 [1.51-5.51] | 1.37E-3 | 2.68 [1.69-4.25] | 2.60E-5 |
| 19 | 46090283 | rs141771937 | T | C | NA | NA | NA | NA | 1.00 | 0.0133 | 2.33 [1.30-4.20] | 4.71E-3 | 0.97 | 0.0125 | 4.31 [2.32-8.01] | 3.87E-6 | 3.08 [1.99-4.76] | 3.86E-7 |
| 19 | 49909139 | rs559050154 | G | A | NA | NA | NA | NA | 0.91 | 0.0115 | 3.64 [1.86-7.14] | 1.63E-4 | 0.88 | 0.0067 | 3.25 [1.09-9.68] | 3.44E-2 | 3.54 [1.98-6.31] | 1.88E-5 |
| 20 | 10720969 | rs78188241 | T | C | 0.82 | 0.0211 | 2.97 [1.58-5.59] | 7.15E-4 | 1.00 | 0.0301 | 1.53 [1.00-2.34] | 4.81E-2 | 0.95 | 0.0212 | 2.55 [1.53-4.25] | 3.35E-4 | 2.05 [1.52-2.76] | 2.11E-6 |
| 20 | 11731982 | rs117414348 | A | G | NA | NA | NA | NA | 0.84 | 0.0111 | 3.30 [1.77-6.17] | 1.81E-4 | 0.80 | 0.0060 | 3.23 [1.30-8.00] | 1.13E-2 | 3.28 [1.95-5.52] | 8.01E-6 |
| 20 | 24359932 | rs77942623 | T | C | 0.82 | 0.0051 | 7.75 [2.07-29.00] | 2.34E-3 | NA | NA | NA | NA | 0.92 | 0.0052 | 6.97 [2.76-17.58] | 3.96E-5 | 7.21 [3.27-15.90] | 9.84E-7 |
| 20 | 36479360 | rs76505427 | G | T | NA | NA | NA | NA | 0.86 | 0.0061 | 3.05 [1.31-7.10] | 9.52E-3 | 0.86 | 0.0074 | 5.58 [2.54-12.3] | 1.92E-5 | 4.16 [2.31-7.50] | 2.03E-6 |
| 20 | 39496129 | rs118012081 | G | A | NA | NA | NA | NA | 0.82 | 0.0071 | 2.80 [1.12-7.01] | 2.77E-2 | 0.82 | 0.0082 | 6.78 [2.93-15.70] | 7.77E-6 | 4.45 [2.37-8.39] | 3.69E-6 |
| 20 | 40341113 | rs142276787 | T | C | 0.88 | 0.0078 | 3.34 [1.15-9.65] | 2.61E-2 | 0.95 | 0.0071 | 2.76 [1.21-6.28] | 1.58E-2 | 0.94 | 0.0059 | 3.15 [1.37-7.25] | 7.03E-3 | 3.02 [1.78-5.12] | 3.89E-5 |
| 20 | 42789152 | rs12480648 | C | T | 0.95 | 0.0063 | 3.99 [1.21-13.10] | 2.28E-2 | 0.94 | 0.0060 | 6.56 [2.41-17.90] | 2.35E-4 | NA | NA | NA | NA | 5.40 [2.47-11.80] | 2.38E-5 |
| 20 | 48957748 | rs76672992 | G | A | 0.83 | 0.0067 | 4.50 [1.59-12.80] | 4.72E-3 | 0.96 | 0.0106 | 2.98 [1.63-5.46] | 3.86E-4 | 0.97 | 0.0127 | 2.78 [1.35-5.73] | 5.62E-3 | 3.11 [2.02-4.79] | 2.82E-7 |

**Table E1**. Stage I variants of nominal significance identified in the 3-way meta-analysis

| Variant Location and Alleles | | | | | US cohort | | | | UK cohort | | | | UUS cohort | | | | Stage-I meta-analysis | |
|---|---|---|---|---|---|---|---|---|---|---|---|---|---|---|---|---|---|---|
| Chr | Pos | SNP rsID | REF | EA | R2 | EAF | HR [95%CI] | P | R2 | EAF | HR [95%CI] | P | R2 | EAF | HR [95%CI] | P | HR [95%CI] | P |
| 20 | 61469020 | rs71325411 | A | G | NA | NA | NA | NA | 0.75 | 0.0335 | 2.32 [1.49-3.61] | 1.96E-4 | 0.74 | 0.0287 | 2.07 [1.20-3.59] | 9.33E-3 | 2.22 [1.57-3.15] | 7.70E-6 |
| 22 | 19257205 | rs362240 | A | G | NA | NA | NA | NA | 0.90 | 0.0093 | 4.04 [1.94-8.41] | 1.86E-4 | 0.89 | 0.0101 | 2.62 [1.35-5.11] | 4.63E-3 | 3.22 [1.94-5.33] | 5.48E-6 |
| 22 | 34904345 | rs187511566 | A | G | NA | NA | NA | NA | 0.90 | 0.0053 | 6.70 [2.71-16.60] | 3.73E-5 | 0.84 | 0.0056 | 3.69 [1.43-9.49] | 6.79E-3 | 5.10 [2.62-9.92] | 1.62E-6 |
| 22 | 36599507 | rs9306307 | A | G | 0.89 | 0.2510 | 0.71 [0.55-0.91] | 7.17E-3 | 0.96 | 0.2905 | 0.78 [0.65-0.93] | 5.93E-3 | 0.97 | 0.2888 | 0.79 [0.65-0.97] | 2.24E-2 | 0.77 [0.68-0.87] | 1.70E-5 |
| 22 | 38095241 | rs79385984 | G | A | NA | NA | NA | NA | 0.82 | 0.0104 | 2.86 [1.47-5.60] | 2.09E-3 | 0.84 | 0.0077 | 3.62 [1.50-8.74] | 4.21E-3 | 3.11 [1.81-5.34] | 4.03E-5 |

| Table E2. Stage II validation of survival-associated variants identified in stage I. | | | | | |
|---|---|---|---|---|---|
| **Chr** | **SNP rs ID** | **Position** | **EAF** | **HR [95%CI]** | ***p*** |
| 1 | rs7514663 | 10762349 | 0.008 | 0.91 [0.23-3.68] | 0.90 |
| 1 | rs185926659 | 94606086 | NA | NA | NA |
| 1 | rs148192333 | 100493151 | 0.004 | 1.09 [0.27-4.44] | 0.90 |
| 1 | rs111749519 | 170154993 | 0.008 | 0.97 [0.40-2.35] | 0.94 |
| 1 | rs190285902 | 188069617 | 0.013 | 1.07 [0.48-2.42] | 0.86 |
| 1 | rs145551873 | 188151083 | 0.016 | 1.08 [0.53-2.19] | 0.84 |
| 1 | rs11589092 | 200768521 | 0.100 | 0.94 [0.70-1.25] | 0.67 |
| 1 | rs968549 | 200861595 | 0.903 | 1.07 [0.80-1.43] | 0.66 |
| 1 | rs115684501 | 203327373 | 0.004 | 1.91 [0.47-7.74] | 0.36 |
| 1 | rs115702239 | 214247966 | 0.008 | 2.17 [0.8-5.84] | 0.13 |
| 1 | rs112891824 | 222319103 | 0.016 | 1.12 [0.57-2.18] | 0.74 |
| 1 | rs34473966 | 242551121 | NA | NA | NA |
| 2 | rs114698213 | 38114198 | 0.008 | 0.47 [0.12-1.88] | 0.28 |
| 2 | rs150622385 | 38249347 | 0.008 | 0.88 [0.32-2.39] | 0.81 |
| 2 | rs185306441 | 43602198 | 0.012 | 1.78 [0.91-3.48] | 0.09 |
| 2 | rs115376562 | 47162883 | 0.010 | 1.24 [0.55-2.80] | 0.61 |
| 2 | rs150765591 | 54643373 | 0.008 | 2.17 [0.96-4.92] | 0.06 |
| 2 | rs184498750 | 84291167 | 0.012 | 2.05 [1.00-4.18] | 0.0493 |
| 2 | rs149029025 | 138337791 | NA | NA | NA |
| 2 | rs142306180 | 142907569 | 0.008 | 1.36 [0.56-3.32] | 0.50 |
| 2 | rs2357323 | 172941078 | 0.004 | 0.71 [0.18-2.86] | 0.63 |
| 2 | rs56923699 | 172942149 | 0.004 | 0.71 [0.18-2.86] | 0.63 |
| 2 | rs182271851 | 173111445 | 0.004 | 0.48 [0.12-1.94] | 0.30 |
| 2 | rs377036225 | 226721440 | 0.009 | 0.41 [0.13-1.29] | 0.13 |
| 2 | rs142266733 | 226758606 | 0.009 | 0.41 [0.13-1.29] | 0.13 |
| 2 | rs188643417 | 240384770 | 0.006 | 0.60 [0.15-2.43] | 0.48 |

| Table E2. Stage II validation of survival-associated variants identified in stage I. | | | | | |
|---|---|---|---|---|---|
| **Chr** | **SNP rs ID** | **Position** | **EAF** | **HR [95%CI]** | ***p*** |
| 2 | rs116377902 | 241253363 | 0.009 | 0.39 [0.10-1.56] | 0.18 |
| 2 | rs75406448 | 241397273 | 0.045 | 1.00 [0.65-1.53] | 0.99 |
| 3 | rs73177699 | 20681518 | 0.004 | 2.85 [0.90-9.04] | 0.08 |
| 3 | rs77895452 | 59911730 | 0.004 | 0.54 [0.13-2.27] | 0.40 |
| 3 | rs139644937 | 69741636 | 0.011 | 0.88 [0.41-1.87] | 0.74 |
| 3 | rs142255892 | 75034442 | 0.012 | 1.12 [0.51-2.42] | 0.78 |
| 3 | rs36022026 | 79693632 | 0.065 | 0.94 [0.66-1.33] | 0.72 |
| 3 | rs2178398 | 112828302 | 0.176 | 0.92 [0.72-1.16] | 0.47 |
| 3 | rs368666349 | 145344805 | 0.007 | 1.22 [0.30-4.94] | 0.78 |
| 4 | rs191600377 | 11752169 | 0.011 | 1.81 [0.80-4.11] | 0.15 |
| 4 | rs17260001 | 11982650 | 0.008 | 1.67 [0.61-4.58] | 0.32 |
| 4 | rs192085162 | 12091599 | NA | NA | NA |
| 4 | rs142199841 | 66572997 | 0.007 | 0.72 [0.22-2.30] | 0.58 |
| 4 | rs79914686 | 74459567 | 0.002 | 3.95 [0.98-16.0] | 0.05 |
| 4 | rs28641522 | 76495474 | 0.007 | 3.1 [0.98-9.82] | 0.05 |
| 4 | rs6850444 | 122351715 | 0.352 | 0.89 [0.74-1.07] | 0.22 |
| 4 | rs71618464 | 166645872 | 0.008 | 1.56 [0.72-3.38] | 0.26 |
| 4 | rs139698405 | 175369612 | 0.009 | 0.57 [0.18-1.80] | 0.34 |
| 4 | rs190563090 | 175401693 | 0.009 | 0.57 [0.18-1.80] | 0.34 |
| 5 | rs138830243 | 36581565 | 0.006 | 1.13 [0.28-4.55] | 0.87 |
| 5 | rs138577491 | 96796936 | 0.010 | 0.48 [0.15-1.58] | 0.23 |
| 5 | rs571176768 | 106109149 | NA | NA | NA |
| 5 | rs191657347 | 151623819 | 0.006 | 1.61 [0.51-5.14] | 0.42 |
| 5 | rs536809954 | 154587673 | 0.009 | 1.00 [0.37-2.70] | 1.00 |
| 5 | rs13172001 | 154820302 | 0.011 | 2.07 [0.97-4.42] | 0.06 |
| 6 | rs116594669 | 26118992 | 0.007 | 0.37 [0.05-2.66] | 0.32 |

| Table E2. Stage II validation of survival-associated variants identified in stage I. | | | | | |
|---|---|---|---|---|---|
| **Chr** | **SNP rs ID** | **Position** | **EAF** | **HR [95%CI]** | ***p*** |
| 6 | rs145442342 | 75389935 | 0.011 | 0.72 [0.29-1.76] | 0.47 |
| 6 | rs140590158 | 77044422 | 0.020 | 0.75 [0.38-1.47] | 0.40 |
| 6 | rs116863455 | 84945776 | 0.011 | 0.87 [0.32-2.35] | 0.79 |
| 6 | rs149057325 | 94466663 | 0.027 | 1.01 [0.60-1.69] | 0.97 |
| 6 | rs143246710 | 94498243 | 0.015 | 0.98 [0.52-1.87] | 0.96 |
| 6 | rs144768626 | 94531687 | 0.015 | 0.99 [0.52-1.88] | 0.97 |
| 6 | rs139568075* | 102917669 | 0.009 | 0.10 [0.01-0.73] | 0.02 |
| 6 | rs35483630 | 116041139 | 0.010 | 0.67 [0.30-1.52] | 0.34 |
| 6 | rs181590625 | 116377677 | 0.008 | 0.63 [0.25-1.57] | 0.32 |
| 6 | rs535860622 | 139248572 | 0.006 | 1.30 [0.32-5.26] | 0.71 |
| 6 | rs149730644 | 143298800 | NA | NA | NA |
| 6 | rs62427038 | 152786447 | 0.003 | 0.86 [0.21-3.48] | 0.83 |
| 6 | rs188589004 | 164521835 | 0.007 | 0.94 [0.30-2.97] | 0.91 |
| 7 | rs183625998 | 1065220 | 0.006 | 0.83 [0.20-3.36] | 0.79 |
| 7 | rs192803195 | 1120020 | 0.007 | 0.81 [0.20-3.28] | 0.77 |
| 7 | rs868964452 | 74953934 | NA | NA | NA |
| 7 | rs117388086 | 75344084 | 0.012 | 0.39 [0.12-1.23] | 0.11 |
| 7 | rs111489307 | 75358307 | 0.016 | 0.64 [0.24-1.73] | 0.38 |
| 7 | rs149406562 | 101163592 | 0.012 | 2.00 [0.82-4.91] | 0.13 |
| 7 | rs142639517 | 103029848 | 0.010 | 1.20 [0.49-2.93] | 0.68 |
| 7 | rs74580469 | 125546344 | 0.010 | 0.33 [0.08-1.33] | 0.12 |
| 7 | rs556475473 | 148106376 | NA | NA | NA |
| 7 | rs73728959 | 152982958 | 0.008 | 0.55 [0.14-2.23] | 0.40 |
| 8 | rs148904289 | 9496015 | 0.010 | 0.86 [0.27-2.70] | 0.80 |
| 8 | rs113465261 | 22381858 | 0.009 | 1.31 [0.58-2.97] | 0.52 |
| 8 | rs193064386 | 23454598 | 0.003 | 2.77 [0.67-11.5] | 0.16 |

| Table E2. Stage II validation of survival-associated variants identified in stage I. | | | | | |
|---|---|---|---|---|---|
| **Chr** | **SNP rs ID** | **Position** | **EAF** | **HR [95%CI]** | ***p*** |
| 8 | rs79562205 | 23626607 | 0.012 | 1.09 [0.48-2.46] | 0.84 |
| 8 | rs116969973 | 80493124 | 0.008 | 1.83 [0.74-4.53] | 0.19 |
| 8 | rs183828688* | 92675281 | 0.007 | 0.38 [1.08-6.5] | 0.03 |
| 8 | rs117931584 | 120287198 | 0.011 | 0.98 [0.4-2.41] | 0.97 |
| 9 | rs147994675 | 4064850 | 0.013 | 1.13 [0.53-2.44] | 0.75 |
| 9 | rs190922348 | 7042171 | 0.010 | 1.54 [0.67-3.50] | 0.31 |
| 9 | rs118165660 | 14343426 | 0.017 | 1.03 [0.53-2.01] | 0.93 |
| 9 | rs113029495 | 24585027 | 0.007 | 0.79 [0.29-2.14] | 0.65 |
| 9 | rs139230397 | 86260066 | 0.003 | 1.77 [0.25-12.8] | 0.57 |
| 9 | rs187088797 | 92489004 | 0.008 | 1.72 [0.64-4.65] | 0.29 |
| 10 | rs142562867 | 4620992 | 0.017 | 0.82 [0.41-1.67] | 0.59 |
| 10 | rs111589468 | 11875431 | 0.006 | 0.70 [0.17-2.84] | 0.62 |
| 10 | rs139494937 | 43302339 | 0.020 | 1.36 [0.74-2.53] | 0.32 |
| 10 | rs186597433 | 73718436 | 0.006 | 2.31 [0.85-6.32] | 0.10 |
| 10 | rs191887805 | 87849922 | 0.007 | 0.75 [0.24-2.34] | 0.62 |
| 10 | rs117054238 | 120582311 | 0.006 | 2.14 [0.79-5.80] | 0.13 |
| 11 | rs78904863 | 14115243 | 0.022 | 0.55 [0.26-1.18] | 0.13 |
| 11 | rs111945608 | 28663945 | 0.019 | 0.56 [0.24-1.3] | 0.18 |
| 11 | rs189350185 | 30638253 | 0.007 | 0.51 [0.16-1.61] | 0.25 |
| 11 | rs76061348 | 33998481 | 0.015 | 0.68 [0.28-1.66] | 0.39 |
| 11 | rs118080683 | 36108905 | 0.029 | 0.57 [0.31-1.06] | 0.07 |
| 11 | rs186991973 | 44113701 | 0.007 | 2.16 [0.85-5.46] | 0.10 |
| 11 | rs186259260 | 75214357 | 0.010 | 1.17 [0.51-2.66] | 0.71 |
| 11 | rs72940008 | 81937642 | 0.013 | 0.63 [0.23-1.72] | 0.37 |
| 11 | rs118091577 | 123294624 | 0.003 | NA | 0.99 |
| 11 | rs559811602 | 123312059 | 0.001 | NA | 0.99 |

| Table E2. Stage II validation of survival-associated variants identified in stage I. | | | | | |
|---|---|---|---|---|---|
| **Chr** | **SNP rs ID** | **Position** | **EAF** | **HR [95%CI]** | ***p*** |
| 11 | rs189162333 | 126441296 | 0.023 | 1.11 [0.62-1.99] | 0.73 |
| 11 | rs192803572 | 132893929 | 0.007 | 0.40 [0.10-1.62] | 0.20 |
| 11 | rs140676527 | 134086356 | 0.004 | 0.40 [0.06-2.85] | 0.36 |
| 12 | rs150598029 | 5522297 | 0.010 | 0.57 [0.21-1.54] | 0.27 |
| 12 | rs185814268 | 6067283 | 0.007 | 1.41 [0.35-5.78] | 0.63 |
| 12 | rs4760248 | 59333262 | NA | NA | NA |
| 12 | rs117273906 | 77326010 | 0.017 | 0.70 [0.29-1.71] | 0.43 |
| 12 | rs61926456 | 91345195 | 0.016 | 1.05 [0.51-2.17] | 0.89 |
| 12 | rs188979614 | 124110116 | 0.001 | NA | 0.99 |
| 12 | rs190737725 | 124161354 | 0.002 | 0.67 [0.09-4.80] | 0.69 |
| 13 | rs192910570 | 43516465 | 0.017 | 0.63 [0.26-1.53] | 0.30 |
| 13 | rs143441960 | 43523271 | 0.020 | 0.72 [0.34-1.54] | 0.40 |
| 13 | rs35389309 | 50380270 | 0.001 | NA | 0.99 |
| 13 | rs149658103 | 77117893 | 0.016 | 1.33 [0.68-2.60] | 0.40 |
| 13 | rs9556131 | 92556834 | 0.097 | 1.20 [0.88-1.64] | 0.25 |
| 14 | rs182533924 | 53451666 | 0.008 | 1.21 [0.45-3.27] | 0.71 |
| 14 | rs187000606 | 54089652 | 0.009 | 0.96 [0.39-2.34] | 0.92 |
| 14 | rs138142698 | 82049712 | 0.002 | 1.13 [0.28-4.58] | 0.87 |
| 14 | rs117705014 | 92700530 | 0.007 | 1.27 [0.47-3.43] | 0.64 |
| 15 | rs60514164 | 76081200 | 0.076 | 1.42 [1.04-1.93] | 0.0256 |
| 15 | rs116962753 | 82159556 | 0.013 | 0.73 [0.30-1.78] | 0.49 |
| 15 | rs35647788 | 101914234 | 0.008 | 3.12 [1.37-7.11] | 0.0067 |
| 16 | rs183941002 | 13757971 | 0.004 | 1.57 [0.58-4.29] | 0.38 |
| 16 | rs28510778 | 24509583 | 0.063 | 0.98 [0.67-1.43] | 0.91 |
| 16 | rs71387661 | 29437123 | NA | NA | NA |
| 16 | rs147189264 | 29599816 | 0.025 | 1.19 [0.66-2.14] | 0.57 |

| Table E2. Stage II validation of survival-associated variants identified in stage I. | | | | | |
|---|---|---|---|---|---|
| **Chr** | **SNP rs ID** | **Position** | **EAF** | **HR [95%CI]** | ***p*** |
| 16 | rs34944462 | 29603553 | 0.025 | 1.19 [0.66-2.14] | 0.56 |
| 16 | rs148521985 | 57317482 | 0.002 | NA | 0.99 |
| 16 | rs144433549 | 65985243 | 0.007 | 0.59 [0.19-1.87] | 0.37 |
| 16 | rs11642140 | 83235650 | 0.057 | 0.85 [0.57-1.27] | 0.43 |
| 17 | rs187631060 | 18860566 | 0.001 | 1.32 [0.18-9.67] | 0.78 |
| 17 | rs11650798 | 32965617 | 0.563 | 1.08 [0.9-1.29] | 0.40 |
| 17 | rs187199661 | 78670545 | 0.007 | 0.70 [0.22-2.21] | 0.55 |
| 17 | rs139401580 | 78806880 | 0.003 | 2.20 [0.54-8.94] | 0.27 |
| 18 | rs187303689 | 12796936 | 0.007 | 0.47 [0.11-1.92] | 0.29 |
| 18 | rs191153148 | 12954976 | 0.007 | 0.38 [0.09-1.57] | 0.18 |
| 18 | rs10469051 | 25516518 | 0.071 | 1.10 [0.76-1.59] | 0.61 |
| 18 | rs141194630 | 30461003 | 0.004 | 1.07 [0.27-4.32] | 0.92 |
| 19 | rs150244663 | 1021639 | 0.021 | 1.04 [0.61-1.8] | 0.88 |
| 19 | rs147560834 | 1412576 | 0.011 | 0.70 [0.29-1.72] | 0.44 |
| 19 | rs3893252 | 1412985 | 0.015 | 2.09 [1.05-4.15] | 0.0357 |
| 19 | rs78359732 | 1482080 | 0.010 | 0.84 [0.34-2.07] | 0.70 |
| 19 | rs141771937 | 46090283 | 0.011 | 0.73 [0.27-1.98] | 0.54 |
| 19 | rs559050154 | 49909139 | 0.003 | 0.82 [0.20-3.33] | 0.78 |
| 20 | rs78188241 | 10720969 | 0.027 | 1.47 [0.85-2.53] | 0.17 |
| 20 | rs117414348 | 11731982 | 0.006 | 0.81 [0.2-3.26] | 0.76 |
| 20 | rs77942623 | 24359932 | 0.004 | 1.69 [0.62-4.61] | 0.31 |
| 20 | rs76505427 | 36479360 | 0.007 | 1.15 [0.28-4.75] | 0.84 |
| 20 | rs118012081 | 39496129 | 0.007 | 0.45 [0.11-1.81] | 0.26 |
| 20 | rs142276787 | 40341113 | 0.010 | 0.85 [0.31-2.32] | 0.75 |
| 20 | rs12480648 | 42789152 | 0.008 | 0.82 [0.30-2.21] | 0.69 |
| 20 | rs76672992 | 48957748 | 0.011 | 1.88 [0.92-3.82] | 0.08 |

| Table E2. Stage II validation of survival-associated variants identified in stage I. | | | | | |
|---|---|---|---|---|---|
| **Chr** | **SNP rs ID** | **Position** | **EAF** | **HR [95%CI]** | ***p*** |
| 20 | rs71325411 | 61469020 | 0.055 | 1.30 [0.92-1.83] | 0.14 |
| 22 | rs362240 | 19257205 | 0.008 | 1.62 [0.66-3.99] | 0.29 |
| 22 | rs187511566 | 34904345 | 0.007 | 1.75 [0.71-4.29] | 0.22 |
| 22 | rs9306307 | 36599507 | NA | NA | NA |
| 22 | rs79385984 | 38095241 | 0.017 | 1.41 [0.72-2.76] | 0.31 |

*p<0.05 but with an opposite direction of effect compared to Stage I

**Table E3. Survival association for top variants before and after multivariable adjustment**

| Variant location (hg19) and nearest gene | | | | | | Unadjusted | | Adjusted model 1* | | Adjusted model 2** | |
|---|---|---|---|---|---|---|---|---|---|---|---|
| **Chr.** | **Position** | **SNP rsID** | **Gene** | **REF** | **EA** | **HR [95% CI]** | ***P-value*** | **HR [95 % CI]** | ***P-value*** | **HR [95 %CI]** | ***P-value*** |
| 2 | 84291167 | rs184498750 | *SUCLG1* | G | T | 3.11 [1.88-5.15] | $9.83x10^{-6}$ | 1.84 (1.13-2.99) | 0.013 | 2.02 (1.24-3.28) | 0.005 |
| 15 | 76081200 | rs60514164 | *UBE2Q2* | C | T | 1.6 [1.32-1.95] | $2.35x10^{-6}$ | 1.48 (1.20-1.84) | $3.11x10^{-4}$ | 1.47 (1.19-1.83) | $3.83x10^{-5}$ |
| 15 | 101914234 | rs35647788 | *PCSK6* | C | T | 4.76 [2.62-8.64] | $2.96x10^{-7}$ | 4.64 (2.38-9.04) | $6.32x10^{-6}$ | 4.18 (2.15-8.15) | $2.45x10^{-5}$ |
| 19 | 1412985 | rs3893252 | *DAZAP1* | C | T | 3.57 [1.97-6.49] | $2.91x10^{-5}$ | 1.39 (0.74-2.61) | 0.304 | 1.29 (0.68-2.44) | 0.432 |

* Adjusted for baseline GAP stage

** Adjusted for sex and baseline age, FVC (% predicted) and DLCO (% predicted)

**Table E4. Characteristics of patients with PCSK6 variant**

| Patient | Study | Centre | Risk Alleles | Outcome | Survival (months) |
|---|---|---|---|---|---|
| 1 | UK | Edinburgh | 1 | Death | 9.63 |
| 2 | UK | Edinburgh | 1 | Death | 38.83 |
| 3 | UK | Edinburgh | 1 | Death | 26.2 |
| 4 | UK | PROFILE | 1 | Death | 5.62 |
| 5 | UK | PROFILE | 1 | Death | 7.92 |
| 6 | UK | Trent Lung | 1 | Death | 38.79 |
| 7 | UK | Trent Lung | 1 | Death | 19.13 |
| 8 | UUS | Brompton | 1 | Death | 10 |
| 9 | UUS | Brompton | 1 | Death | 8 |
| 10 | UUS | Brompton | 1 | Death | 2 |
| 11 | UUS | Brompton | 1 | Alive | 10 |
| 12 | UUS | Chicago | 1 | Alive | 1.18 |
| 13 | UUS | Chicago | 1 | Alive | 16.79 |
| 14 | UUS | Chicago | 1 | Alive | 5.36 |
| 15 | UUS | Chicago | 1 | Death | 20.4 |
| 16 | UUS | Chicago | 1 | Alive | 7.43 |
| 17 | UUS | Nottingham | 1 | Death | 6.44 |
| 18 | UUS | UCD | 1 | Death | 11.89 |
| 19 | UUS | UCD | 1 | Alive | 22.8 |

**Table E5. Sensitivity analysis comparing PCSK6 results with censoring lung transplant versus considering this an event**

| | UK | | UUS | | Meta | |
|---|---|---|---|---|---|---|
| | OR [95% CI] | p | OR [95% CI] | p | OR [95% CI] | p |
| **Transplant as event** | 5.18 [2.35, 11.42] | 4.60E-05 | 4.25 [1.78, 10.17] | 1.14E-03 | 4.76 [2.62, 8.64] | 2.96E-07 |
| **Transplant censored** | 5.23 [2.41, 11.35] | 2.83E-05 | 4.48 [1.87, 10.74] | 7.67E-04 | 4.89 [2.73, 8.73] | 8.04E-08 |

| **Table E6.** *In silico* functional assessment variants associated with TFS after stage II | | | | |
|---|---|---|---|---|
| | **rs184498750** *(SUCLG1)* | **rs60514164** *(UBE2Q2)* | **rs35647788** *(PCSK6)* | **rs3893252** *(DAZAP1)* |
| **RegulomedB rank (Score)** | 5 - TF binding or Dnase peak (0) | 5 - TF binding or Dnase peak (0.17) | 4 - TF binding + Dnase peak (0.61) | 3a - TF binding + any motif + DNase peak (0.73) |
| **Enhancer histone marks** [HaploReg] | None | H3K4me1 (Epithelial) | H3K4me1[a], H3K27ac[b] | H3K4me1[c], H3K27ac[d] |
| **Promoter histone marks** [HaploReg] | H3K9ac (Fetal lung) | None | H3K4me3[e], H3K9ac[f] | H3K4me3[g], H3K9ac[h] |
| **DNAse** [HaploReg] | None | None | Digestive | HSC & B-cell, Muscle, Fetal Adrenal Gland, Pancreas, Monocytes-CD14+ RO01746 Primary Cells |
| **Altered regulatory motifs** [HaploReg] | CIZ, Mef2 | AIRE_2, AP-3 | ZBTB33 | None |
| **Proteins bound** [HaploReg] | None | None | None | None |
| **CHICP**<br>[Cell line: *gene* (SCORE)].<br>A default score of 5 was considered as threshold to identify PCHi-C interactions | None | GM12878: *SIN3A* (10.01), *MAN2C1* (9.56), *NRG4* and *C15ORF27* (9.33) | CD34: *TM2D3* (9.90); GM12878: *TM2D3* (8.65), *OR4F6* (9.48), *OR4F15* (9.64), *CHSY1* (8.04) | Neutrophils: BSG (8.46), POLRMT (5.46); CD34: DAZAP1 and RPS15 (12.99); GM12878: MUM and EFNA2 (10.87); CIRBP and C19orf24 (11.40); C19orf26, ATP5D, MIDN, and CIRBP (10.90); SBNO2 and STK11 (10.41); HMHA1 (10.17); WDR18, GRIN3B, and TMEM259 (10.52); PTBP1, LPPR3, and AZU1 (9.68); CSNK1G2 (9.16); KLF16 (10.40); ATP8B3 and REXO1 (10.03); ONECUT3 (10.75); TCF3 (Scores 9.99 and 10.21); REEP6, PCSK4, and C19orf25 (11.65); C19orf25 and APC2 (12.38); DAZAP1 and RPS15 (12.95); hESC Derived Cardiomyocytes: CIRBP, CIRBP-AS1 and C19orf24 (6.59); MUM1 and EFNA2 (5.69) |
| **Open Targets Genetics**<br>Top ranked genes based on the overall V2G score | *AC106874.1* (top ranked), *SUCLG1*, *DNAH6* | *SNUPN* (top ranked), *MAN2C1*, *AC105020.1*, *IMP3* | *PCSK6* (top ranked), *PCSK6-AS1*, *SELENOS*, *SNRPA1* | DAZAP1 (top ranked), NDUFS7, AC005329.2 |
| **dsQTL [GTEx]**<br>Tissue-specific $P \leq 0.05$ | None | *MAN2C1* (Adipose, skin, artery, esophagus) | None | None |
| **eQTL [GTEx]**<br>Tissue-specific $P \leq 0.05$ | None | *MAN2C1* (lung, cultured fibroblasts, adipose, skin, artery, esophagus, among others) | None | *LLNLR-307A6.1* (skin) |
| **Score CAPE dsQTL >0.5 [SNPDelScore]** | NA | None | NA | H1 BMP4 derived trophoblast cultured cells, IMR90 fetal lung fibroblasts cell line, A549 EtOH 0.02pct lung carcinoma cell line, GM12878 lymphoblastoid cells, HeLa-S3 Cervical carcinoma cell line, Monocytes-CD14+ RO01746 Primary cells, NH-A Astrocytes primary cells, NHEK-Epidermal keratinocyte primary cells |

| **Table E6.** *In silico* functional assessment variants associated with TFS after stage II | | | | |
|---|---|---|---|---|
| | **rs184498750** *(SUCLG1)* | **rs60514164** *(UBE2Q2)* | **rs35647788** *(PCSK6)* | **rs3893252** *(DAZAP1)* |
| **Score CAPE eQTL >0.5 [SNPDelScore]** | NA | iPS DF 6.9 Cells, primary T cells from peripheral blood, foreskin fibroblast primary cells, gastric, ovary, GM12878 lymphoblastoid cells, HSMM skeletal muscle myoblasts cells, HSMM cell derived skeletal muscle myotubes cells, NH-A Astrocytes primary cells | NA | H1 cells, H9 cells, iPS DF 6.9 Cells, iPS DF 19.11 Cells, fetal intestine small, fetal muscle leg, placenta, ovary, HMEC mammary epithelial primary cells, HSMM skeletal muscle myoblasts cells, HSMM cell derived skeletal muscle myotubes cells, NHDF-Ad Adult Dermal Fibroblast primary cells |
| **PheWAS (PhenoScanner)** | Allele T has positive/increased effect association with:<br>-Cause of death: hodgkins disease, unspecified (*P*=$1.51\times10^{-6}$)<br>-Self-reported gall bladder disease (*P*=$1.50\times10^{-5}$)<br>-Cause of death: ischaemic cardiomyopathy (*P*=$3.39\times10^{-4}$)<br>- Treatment with dexamethasone (*P*=$3.65\times10^{-4}$) | Allele T has positive/increased effect association with:<br>-Self-reported nasal or sinus disorder (*P*=$3.98\times10^{-5}$)<br>-Zoster (*P*=$2.17\times10^{-4}$)<br>-Treatment with logynon tablet (*P*=$6.84\times10^{-4}$)<br>-Duration of other exercises (*P*=$9.17\times10^{-4}$)<br><br>Allele T has negative/decreased effect association with:<br>- Treatment with becotide 50 inhaler (*P*=$5.84\times10^{-4}$) | Allele T has positive/increased effect association with:<br>-Self-reported fracture lower leg or ankle (*P*=$4.39\times10^{-5}$)<br>-Self-reported acute infective polyneuritis or Guillain-Barre syndrome (*P*=$1.16\times10^{-4}$)<br>-Self-reported multiple sclerosis (*P*=$2.27\times10^{-4}$)<br>-Cause of death: epilepsy, unspecified (*P*=$3.61\times10^{-4}$)<br>-Polyarthrosis (*P*=$5.07\times10^{-4}$) | Allele T has positive/increased effect association with:<br>-Home area population density: Scotland large urban area (*P*=$5.09\times10^{-5}$)<br>-Self-reported unclassifiable non-cancer illness (*P*=$1.07\times10^{-4}$)<br>-Cause of death: duodenum (*P*=$1.24\times10^{-4}$)<br>-Self-reported thyroid problem (*P*=$1.74\times10^{-4}$)<br>-Cause of death: tongue, unspecified (*P*=$2.70\times10^{-4}$)<br>-Cause of death: vascular dementia, unspecified (*P*=$3.18\times10^{-4}$)<br>- Benign neoplasm of major salivary glands (*P*=$6.06\times10^{-4}$) |

CAPE, cellular dependent deactivating mutations; CD34, human hematopoietic progenitor cell line; CHICP, Capture HiC Plotter; dsQTL, DNase I-sensitive quantitative trait loci; eQTL, expression quantitative trait loci; ESC, embryonic stem cells; GM12878, lymphoblastoid cell line; HSC, hematopoietic stem cells; HSMM, human skeletal muscle myoblasts; HUVEC, human umbilical vein endothelial cell; iPS DF 6.9 Cells, human induced pluripotent stem cell line derived from foreskin fibroblasts; IMR90, human fetal lung cells; iPSC, induced pluripotent stem cells; LD, linkage disequilibrium; NH-A, normal human Astrocytes; NHDF, normal human dermal fibroblasts; NHLF, normal human lung fibroblasts; PCHi-C, Promoter Capture Hi-C; TF, transcription factor; V2G, variants to genes.

[a]ESC, iPSC, ES-derive, Blood & T-cell, HSC & B-cell, Epithelial (Foreskin Keratinocyte Primary Cells skin03), Brain, Muscle, Heart, Digestive, Fetal Adrenal Gland, Liver, Pancreas, Lung, Spleen; [b]ES-derive, Brain, Digestive, Liver; [c]IMR90, ESC, iPSC, ES-derive, Blood & T-cell, HSC & B-cell, Mesench, Myosat, Epithelial, Neurosph, Thymus, Brain, Adipose, Muscle, Heart, Sm. Muscle, Digestive, Placenta Amnion, Fetal Lung, Ovary, Fetal Adrenal Gland, Placenta, Liver, Pancreas, Lung, Spleen, A549 EtOH 0.02pct Lung Carcinoma Cell Line, Dnd41 T Cell Leukemia Cell Line, GM12878 Lymphoblastoid Cells, HeLa-S3 Cervical Carcinoma Cell Line, HepG2 Hepatocellular Carcinoma Cell Line, HMEC Mammary Epithelial Primary Cells, HSMM Skeletal Muscle Myoblasts Cells, HUVEC Umbilical Vein Endothelial Primary Cells, K562 Leukemia Cells, NH-A Astrocytes Primary Cells, NHEK-Epidermal Keratinocyte Primary Cells, NHLF Lung Fibroblast Primary Cells, Osteoblast Primary Cells; [d]ESC, iPSC, ES-derive, Blood & T-cell, HSC & B-cell, Mesench, Epithelial, Thymus, Brain, Adipose, Muscle, Heart, Sm. Muscle, Digestive, Ovary, Pancreatic Islets, Fetal Adrenal Gland, Placenta, Liver, Pancreas, Lung, Spleen, Dnd41 T Cell Leukemia Cell Line, GM12878 Lymphoblastoid Cells, HeLa-S3 Cervical Carcinoma Cell Line, HUVEC Umbilical Vein Endothelial Primary Cells, K562 Leukemia Cells,Monocytes-CD14+ RO01746 Primary Cells, NH-A Astrocytes Primary Cells, NHDF-Ad Adult Dermal Fibroblast Primary Cells, NHLF Lung Fibroblast Primary Cells, Osteoblast Primary Cells; [e]iPSC, Digestive; [f]Blood & T-cell, Epithelial, Digestive; [g]iPSC, ES-derive, Blood & T-cell, HSC & B-cell, Mesench, Epithelial, Neurosphil, Brain, Adipose, Muscle, Heart, Sm. Muscle, Digestive, Placenta Amnion, Pancreas, Spleen, Dnd41 T Cell Leukemia Cell Line, GM12878 Lymphoblastoid Cells, HeLa-S3 Cervical Carcinoma Cell Line, HepG2 Hepatocellular Carcinoma Cell Line, Monocytes-CD14+ RO01746 Primary Cells, NHLF Lung Fibroblast Primary Cells, Osteoblast Primary Cells; [h]IMR90, ESC, iPSC, ES-derive, Blood & T-cell, Mesench, Epithelial, Brain, Adipose, Muscle, Heart, Sm. Muscle, Digestive, Fetal Lung, Liver, Dnd41 T Cell Leukemia Cell Line, HeLa-S3 Cervical Carcinoma Cell Line, HepG2 Hepatocellular Carcinoma Cell Line, Monocytes-CD14+ RO01746 Primary Cells, NH-A Astrocytes Primary Cells, NHEK-Epidermal Keratinocyte Primary Cells, NHLF Lung Fibroblast Primary Cells

**Table E7. Survival association for fifteen genetic variants previously linked to IPF susceptibility**

| | | | | | | | Risk GWAS | | Survival GWAS | |
|---|---|---|---|---|---|---|---|---|---|---|
| **Chr.** | **Position** | **SNP rsid** | **Locus** | **Ref. allele** | **Effect allele** | **EAF** | **OR* [95% CI]** | ***P-value*** | **HR [95% CI]** | ***P-value*** |
| 3 | 44902386 | rs78238620 | *KIF15* | T | A | 0.08 | 1.58 [1.37, 1.83] | $5.12\times10^{-10}$ | 0.92 [0.76, 1.13] | 0.444 |
| 3 | 169481271 | rs12696304 | *LRRC34 /TERC* | C | G | 0.31 | 1.31 [1.21, 1.40] | $7.09\times10^{-13}$ | 0.95 [0.86, 1.06] | 0.343 |
| 4 | 89885086 | rs2013701 | *FAM13A* | T | G | 0.575 | 1.28 [1.19, 1.35] | $3.30\times10^{-13}$ | 1.01 [0.92, 1.12] | 0.792 |
| 5 | 1282414 | rs7725218 | *TERT* | A | G | 0.734 | 1.39 [1.30, 1.49] | $1.54\times10^{-20}$ | 0.97 [0.86, 1.09] | 0.622 |
| 5 | 169015479 | rs1164837831 | *SPDL1* | G | A | 0.013 | 2.40 [1.70-3.40] | $7.55x10^{-7}$ | 1.23 [0.81-1.87] | 0.327 |
| 6 | 7563232 | rs2076295 | *DSP* | T | G | 0.573 | 1.46 [1.37, 1.56] | $2.79\times10^{-30}$ | 1.07 [0.97, 1.18] | 0.153 |
| 7 | 1909479 | rs12699415 | *MAD1L1* | G | A | 0.469 | 1.28 [1.19, 1.37] | $7.15\times10^{-13}$ | 0.98 [0.89, 1.09] | 0.765 |
| 7 | 99630342 | rs2897075 | 7q22.1 | C | T | 0.408 | 1.30 [1.21, 1.38] | $3.10\times10^{-14}$ | 1.08 [0.98, 1.20] | 0.127 |
| 8 | 120934126 | rs28513081 | *DEPTOR* | G | A | 0.631 | 1.22 [1.15, 1.32] | $1.20\times10^{-9}$ | 0.97 [0.88, 1.08] | 0.559 |
| 11 | 1241221 | rs35705950 | *MUC5B* | G | T | 0.321 | 4.84 [4.37, 5.36] | $1.18\times10^{-203}$ | 0.78 [0.64, 0.94] | 0.008 |
| 13 | 113534984 | rs9577395 | *ATP11A* | G | C | 0.827 | 1.30 [1.20, 1.41] | $1.34\times10^{-10}$ | 0.98 [0.85, 1.11] | 0.723 |
| 15 | 40720542 | rs59424629 | *IVD* | G | T | 0.573 | 1.30 [1.22, 1.41] | $7.30\times10^{-16}$ | 1.12 [1.01, 1.23] | 0.028 |
| 15 | 86097216 | rs62023891 | *AKAP13* | G | A | 0.343 | 1.27 [1.18, 1.36] | $1.27\times10^{-10}$ | 0.98 [0.88, 1.09] | 0.655 |
| 17 | 44214888 | rs2077551 | *MAPT* | C | T | 0.828 | 1.41 [1.30, 1.54] | $2.83\times10^{-16}$ | 0.97 [0.84, 1.12] | 0.675 |
| 19 | 4717672 | rs12610495 | *DPP9* | A | G | 0.371 | 1.31 [1.22, 1.42] | $2.92\times10^{-12}$ | 0.99 [0.87, 1.11] | 0.394 |

Abbreviations: EAF = effect allele frequency; OR = odds ratio; HR = hazard ratio; CI = confidence interval
*Odds ratios are given with respect to the allele that is associated with increased disease risk

**Table E8. Risk association for the four genetic variants linked to IPF survival in the stage I**

| | | | | | | | Risk GWAS | |
|---|---|---|---|---|---|---|---|---|
| **Chr.** | **Position** | **SNP rsid** | **Locus** | **Ref. allele** | **Effect allele** | **EAF** | **OR* [95% CI]** | ***P-value*** |
| 2 | 84291167 | rs184498750 | *SUCLG1* | G | T | 0.01 | 0.90 [0.66, 1.24] | 0.531 |
| 15 | 76081200 | rs60514164 | *UBE2Q2* | C | T | 0.07 | 0.93 [0.82, 1.06] | 0.298 |
| 15 | 101914234 | rs35647788 | *PCSK6* | C | T | 0.01 | 1.01 [0.67, 1.53] | 0.947 |
| 19 | 1412985 | rs3893252 | *DAZAP1* | C | T | 0.01 | 0.74 [0.50, 1.11] | 0.146 |

Abbreviations: EAF = effect allele frequency; OR = odds ratio; HR = hazard ratio; CI = confidence interval
*Odds ratios are given with respect to the allele that is associated with increased disease risk
